# Genotype-phenotype association study conducted on LARGE-PD reveals novel loci associated with Parkinson’s Disease

**DOI:** 10.1101/2025.07.18.25331793

**Authors:** Thiago P Leal, Emily Waldo, Felipe Duarte-Zambrano, Miguel Inca-Martinez, Janvi Ramchandra, Henry Mauricio Chaparro-Solano, Anna E Anello, Victor Borda, Mateus Henrique Gouveia, Daniel Teixeira-dos-Santos, Paula Reyes-Pérez, Emilia Mabel Gatto, Bruno Lopes Santos-Lobato, Gracivane Eufraseo, Grace Helena Letro, Gonzalo Arboleda, Oscar Bernal-Pacheco, Jorge L Orozco, Beatriz Munoz, Pedro Chana-Cuevas, David Aguillon, Sonia Moreno, Gabriel Torrealba-Acosta, Tanya Lobo-Prada, Valentina Muller, C Matias Lopez Razquin, Pedro Braga-Neto, Reyna M Durón, Mayela Rodríguez-Violante, Ana Jimena Hernández-Medrano, Amin Cervantes-Arriaga, Daniel Martinez-Ramirez, Artur F S Schuh, Carlos Roberto de Mello Rieder, Mario Cornejo-Olivas, Julia Rios-Pinto, Angel C Medina, Ivan Cornejo-Herrera, Koni Mejia-Rojas, Angel Vinuela, Vitor Tumas, Angela Vieira Pimentel, Vanderci Borges, Cesar L Avila, Patricio Olguin, Alicia Colombo, Juan Cristobal Nuñez, Alejandra Medina-Rivera, Alejandra E. Ruiz-Contreras, Sarael Alcauter, Elena Dieguez, Karen Nuytemans, Global Parkinson’s Genetics Program, Ignacio F Mata, the Latin American Research Consortium on the Genetics of Parkinson’s Disease (LARGE-PD)

## Abstract

**Background:** The Latin American Research Consortium on the Genetics of Parkinson’s Disease (LARGE-PD) is a multicenter collaboration aimed at understanding the genetic architecture of Parkinson’s disease (PD) in this underrepresented population using data from 15 countries across the Americas and the Caribbean. In this study, we conducted the largest genome-wide association studies (GWAS) for PD susceptibility in Latin Americans.

**Methods:** We analyzed genotype data from LARGE-PD Phase 1 (n = 1,498) and Phase 2 (n = 4,401) using multiple GWAS approaches: SAIGE, which incorporates a genetic relationship matrix in the model; ATT, which includes global ancestry on the model; TRACTOR, which splits allele dosages by ancestry to detect ancestry-specific risk loci; and admixture mapping. We also assessed linkage disequilibrium (LD) patterns and performed Meta-Regression of Multi-AncEstry Genetic Association (MR-MEGA), integrating data from both LARGE-PD phases and two South Asian GWAS.

**Results:** We identified PD-associated loci on chromosomes 1 and 4. Our results replicated previous findings, including the well-established *SNCA* variant rs356182-A (OR = 1.517, p = 1.62×10^−16^). Notably, we identified a locus in *ITPKB* (rs117185933-A, OR = 1.75, p = 3.8×10^−12^), which had the highest CADD Phred score (17.92, top ∼3% most deleterious) among all candidate variants, suggesting strong functional relevance. Functional annotation predicted that this variant may create a premature start codon in the 5′ UTR of *ITPKB*. Although rs117185933-A is in high LD (r^2^ > 0.8) with a variant previously reported by Kishore et al., our LD analysis and MR-MEGA results indicate that this signal is correlated with ancestry heterogeneity and likely represents an independent PD risk locus and a novel putative causal variant. This variant is most frequent in Peruvians from the 1000 Genomes Project (MAF = 0.20) and more common in admixed American populations in gnomAD (MAF = 0.0835), but nearly absent in non-Finnish Europeans (MAF = 0.0002).

**Conclusion:** We identified PD-associated variants in SNCA and ITPKB, the latter not previously reported in European-ancestry studies. The ITPKB variant may lead to a start codon gain in a gene with known protective effects against α-synuclein aggregation *in vivo* and *in vitro* models. These findings underscore the critical importance of including underrepresented populations in genetic research to uncover ancestry-specific risk loci and advance precision medicine for Parkinson’s disease.

## Introduction

Studies indicate that more than 8.5 million people live with Parkinson’s disease (PD) worldwide, and this number could rise to 25.2 million by 2050, with Latin America being one of the populations experiencing the highest increase^1–4^. Genetics plays a crucial role in PD etiology. Genome-wide association studies (GWAS) and GWAS meta-analyses identified ∼130 variants associated with PD with an estimated heritability of up to 28% depending on the studied population^5–9^. Despite these advances, most of our knowledge about PD genetics is based on studies conducted with individuals of European or Asian ancestry, limiting understanding of the genetic architectures of PD in other ancestral populations ^10^.

This lack of genetic diversity poses a significant barrier to genetic diagnosis and treatment, as different populations may have other risk loci and/or tag SNPs^11^. An African GWAS, for example, conducted by researchers from the Global Parkinson’s Genetics Program (GP2) identified a novel PD risk variant in the *GBA1* gene (rs3115534-G), which is relatively common within African ancestry populations but rare in other ancestries^12^.

Addressing these issues, the Latin American Research Consortium on the Genetics of PD (LARGE-PD) was established. This ongoing effort involves over 45 institutions across 15 American and Caribbean countries. LARGE-PD Phase 1 (P1) explored the genetics of PD by genotyping ∼1,500 samples from five South American countries. This dataset was previously used in the first Latin American PD GWAS, as well as in genome-wide analyses of copy number variation, X-chromosome-wide association (XWAS), and polygenic risk score (PRS) inferences based on European GWAS summary statistics^13–16^.

LARGE-PD Phase 2 (P2) is dedicated to advancing the understanding of PD within the Latin American and Caribbean (LAC) populations. This work involves recruiting and genotyping additional samples to use as replication cohorts and increasing the sample size to identify new susceptibility loci for developing PD in Latin American populations. In this study, we performed a comprehensive association analysis, including different GWAS approach and admixture mapping, of PD susceptibility in Latin American populations, using data from LARGE-PD P1 and P2.

## Methods

### Samples

#### Discovery cohort: LARGE-PD Phase 1

We analyzed data from 1,498 individuals (∼53.9% cases) enrolled in the LARGE-PD P1, recruited from Brazil, Chile, Colombia, Peru, and Uruguay, genotyped with the Multi-Ethnic Genotyping Array (Illumina Inc., Chicago, IL, USA). After exclusion of 17 individuals due to missing data, the final dataset included 798 PD cases and 683 controls. A detailed description of the LARGE-PD P1 cohort has been previously reported^14^.

#### Replication cohort: LARGE-PD Phase 2

The 4,401 samples from LARGE-PD P2 were collected between 2019 and 2025 and genotyped using the NeuroBooster Array (NBA) from Illumina. The NBA includes ∼2M variants from Infinium Global Diversity Array and ∼95k variants associated with more than 70 neurological conditions or traits^17^.

Our data includes samples from 10 countries, enrolled across 23 sites. All PD patients were assessed by a local movement disorders specialist and diagnosed using the UK PD Society Brain Bank and/or MDS Clinical Diagnostic Criteria^18,19^. Controls were individuals without major neurological conditions. All participants provided written informed consent according to their respective regional IRB requirements. Additional cohort details are available in the Supporting Information (SI) Section Cohort description (Table S1).

The LARGE-PD P1 and P2 datasets were genotyped with different arrays, so we performed all analyses independently to keep the maximum number of variants and ensure the independence of both datasets. We re-ran the analysis on LARGE-PD P1 to eliminate any potential biases introduced by methodological differences between this study and the approach used by Loesch *et al.*^14^.

### Genotyping Calling and Genetic Quality Control

#### Genotyping calling and quality control

The Illumina signal intensity data files from LARGE-PD P2 were converted to PLINK format using the Illumina Array Analysis Platform (v1.1) using the GP2 cluster file and default parameters. Subsequently, we removed unaligned and problematic variants (see SI section Genotyping Calling).

We applied our quality control (QC) pipeline to LARGE-PD datasets. This pipeline is focused on admixed populations. The pipeline removes individuals and variants based on the following criteria: (i) individuals missing sex and/or status, (ii) individuals with discrepancies between declared and genetic sex (calculated using PLINK v1.90^20^), (iii) ambiguous genotypes, (iv) individuals and variants with more than 5% missing data, (v) duplicated variants (keeping the variant with lower missing rate), (vi) samples with heterozygosity rate ±3 standard deviation (SD) from the mean, (vii) variants failing Hardy-Weinberg Equilibrium (HWE) in controls (p < 1e-6) and cases (p < 1e-10). This process resulted in the related QCed dataset, which was imputed using the TOPMed imputation server panel r3^21^. The number of samples and SNPs is described in Table S2

We generated the list of samples to be removed due to genetic relationships from the QCed dataset using NAToRA^22^ selecting the optimal mode that guarantees minimal sample loss (see SI section on relationship control). All QC analyses were performed using PLINK v2.00a5.10LM^23^, except where a specific tool was mentioned.

### Ancestry analysis

#### Parental Reference dataset

The ancestry analyses were performed using a reference panel of non-admixed samples from the 1000 Genomes Project (1KGP)^24^. The reference panel includes non-admixed samples from African (AFR), East Asian (EAS), European (EUR), Native American (NAT), and South Asian (SAS) metapopulations^25^ (see SI section Ancestry Analysis).

#### Principal Component Analysis

In this study, we employed three approaches of Principal Component Analysis (PCA). For association analyses using all samples, we performed unsupervised PCA on the full dataset to be used as covariates (Figures S1–S4). For analyses incorporating ancestry into the statistical model, we first conducted a supervised PCA using the reference dataset to identify ancestry outliers (Figures S5–S8). After excluding ancestry outliers (samples more than ±3 SD from the mean in the first 10 PCs) and related individuals, we created the unrelated and non-outlier (UNO) dataset. We then performed unsupervised PCA on the UNO dataset to be used as covariates (Figures S9–S12) (see SI section Ancestry Analysis).

#### Ancestry inferences

In genetic studies, ancestry inference refers to the inference of the contribution of each parental population to an individual. In this work, we performed global ancestry (GA) and local ancestry (LA) inference. GA estimates the proportion of genetic ancestry from each ancestral population in an individual. LA identifies the ancestral population of chromosomal segments across an individual’s genome.

GA was inferred using supervised ADMIXTURE^26^, with the previously described 1KGP panel as the reference. LA was inferred using G-nomix [32]. We conducted tests to determine the optimal parameters for performing LA analysis. Our analyses were performed using a three-way admixed reference panel, consisting of EUR and AFR from 1KGP, and a NAT reference panel composed of 1KGP NAT samples and non-admixed controls from LARGE-PD (see SI Section Ancestry Analysis).

### Association studies

#### Genome-Wide Association Studies

Conducting a GWAS in an admixed population presents unique challenges. The genetic heterogeneity within such populations can reduce statistical power and increase susceptibility to false positives and false negatives. In this work, we performed GWAS using three methods: SAIGE^27^, ATT^28^ , and TRACTOR^29^.

SAIGE (Scalable and Accurate Implementation of Generalized mixed model) controls for population stratification and relatedness, incorporating the genetic relationship matrix (GRM), allowing all samples to be included. ATT is a Cochran–Armitage trend test that corrects for GA. TRACTOR is a method that splits the dosage based on LA, assuming the possibility of independent genetic effects across different ancestries.

TRACTOR and SAIGE were selected based on a study^30^ demonstrating that SAIGE was the best method to control the type 1 error rate in admixed cohorts. At the same time, TRACTOR was the best model for identifying ancestry-specific risk loci. ATT was chosen because it exhibits greater statistical power than TRACTOR when the risk allele is common across ancestries^31^.

For SAIGE, we used all samples with age, sex, and the first 10 PCs (Figures S1–S4) as covariates. For TRACTOR and ATT, we used the UNO dataset with sex, age and the 10 PCs (Figures S9–S12) from UNO dataset as covariates.

### Meta-analysis

After performing GWAS, we meta-analyzed data from LARGE-PD P1 and P2 for each method using GWAMA (v2.2.2)^32^ and Meta-Regression of Multi-AncEstry Genetic Association (MR-MEGA) with both phases of LARGE-PD and two SAS GWAS^33,34^. In both we performed under random and fixed-effect models.

While the fixed-effect model assumes one true effect size across studies, the random-effects model assumes that true effect sizes can vary between studies, computing more conservative p-values and wider confidence intervals when heterogeneity is present^35^. Considering the admixture in LAC populations, the random-effects model is more suitable. Finally, heterogeneity was assessed using Cochrane Q statistics and I^2^ calculations (SI section Meta-Analysis).

### Replication of previously identified PD risk loci

To assess the consistency between our findings and previous results, we extracted the beta and p-value from associations reported in Kim et al. 2024 (KIM) and Nalls et al. 2019 (NALLS)^7,8^ studies(GWAS Catalog accession IDs: GCST90275127 and GCST009325) and compared them with the results from the LARGE-PD meta-analysis (SAIGE, ATT, and TRACTOR-NAT random effects). We also selected variants that achieved a suggestive p-value in any LARGE-PD meta-analysis and searched for the reported p-values from both studies.

### Case-control admixture mapping

Case-control admixture mapping (AM) is a LA-phenotype association study that tests the association between LA windows and the phenotype. Since AM involves fewer independent tests than a GWAS, it requires a less stringent p-value threshold^36^. We performed the AM using GENESIS package^37^ using sex, age, and the first 10 PCs as covariates. After this, we extracted variants within the statistically significant regions, as well as those 1 MB flanking these regions, from the UNO dataset and performed association studies using PLINK2 using the same covariates used on AM. We considered statistically significant if p < 1.673x10^−5^ (see SI section Admixture Mapping).

### Functional annotation analysis

After the meta-analysis, we uploaded the summary statistics to the Functional Mapping and Annotation (FUMA) portal^38^. Using FUMA, we identified independent and lead signals within the genomic risk loci based on double clumping criteria. All the analyses used the 1KGP Phase 3 AMR panel as the reference population. Furthermore, MAGMA (Multi-marker Analysis of GenoMic Annotation)^39^ function was used for gene-set analysis using ontology terms from the MSigDB v7.0^40^, setting the default parameters used by FUMA.

Additionally, we investigated the functional impact of the identified variants using the Combined Annotation Dependent Depletion (CADD) score^41^, FORGEdb^42^ score, and SnpEff^43^. While the CADD Phred score estimates the variant’s potential deleteriousness, SnpEff helps interpret the biological impact. The FORGEdb score was created to identify candidate regulatory variants for functional experiments (see SI section Functional annotation).

### Gene expression studies

After obtaining the list with lead and independent variants, we proceeded with the identification of expression quantitative trait loci (eQTLs) using the Genotype-Tissue Expression (GTEx) portal^44^. We compiled a list of replicated variants and queried their rsIDs on the GTEx portal. All genes returned under the “Single-Tissue eQTLs” section were added to our list of candidate genes.

## Results

### Cohorts Description and Ancestry Characterization

P1 includes individuals from five South American countries. P2 expands the cohort to include additional samples from South, Central, and North America, encompassing participants from a total of ten countries (Table 1, Table S1). In terms of demographic composition, the P1 cohort comprises 797 cases (46.9% females) and 681 control (66.4% females). The P2 cohort includes 2,744 cases (42.8% females) and 1,559 controls (67.7% females). The mean age of cases is higher than controls on both phases. The females also had a higher mean age of onset on P2 (56.63 on females vs 55.71 on males).

Our ancestry analysis showed that our samples are primarily 3-way admixed, with contributions of Africans, Europeans, and Native Americans. Considering the minimal contribution of Asian ancestries (Figure 1, Figures S1–S4), our analyses were conducted using a three-way admixed model. To reduce potential bias, ancestry-informed methods were applied after removing outliers identified through projected PCA, thereby preventing misclassification of Asian ancestries in our dataset.

**Figure 1:**
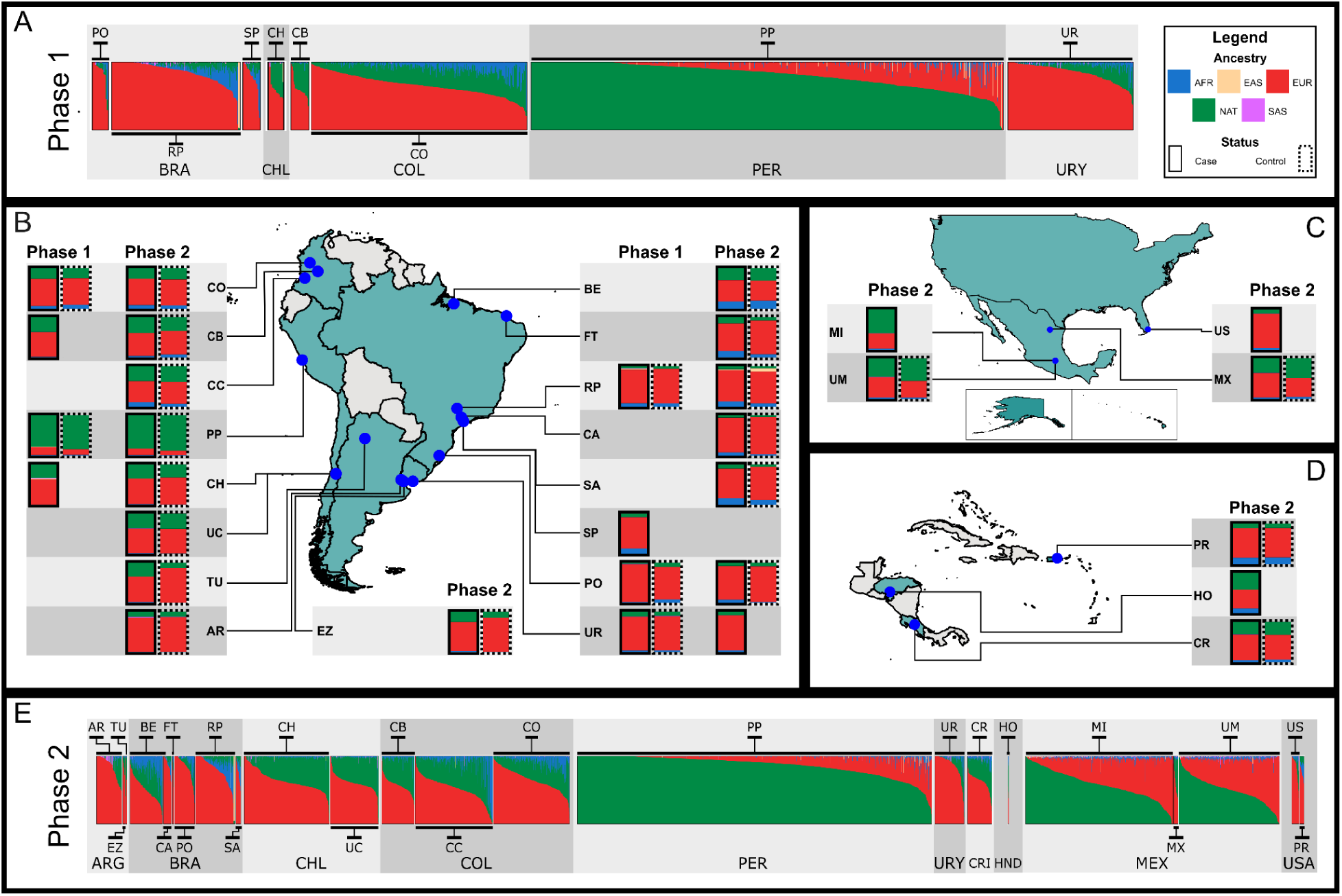
Global ancestry analysis of LARGE-PD Phase 2 using supervised ADMIXTURE. (A) Bar plot for LARGE-PD Phase 1 with the proportions of African (AFR), European (EUR), East Asian (EAS), South Asian (SAS), and Native American (NAT) ancestry clusters. Percentages of continental ancestry stratified by cases (solid lines) and controls (dashed lines) for samples from Phase 1 and 2 for (B) LARGE-PD South America, (C) LARGE-PD North America, (D) LARGE-PD Central America and Caribbean. Arrows on the map indicate the geographic locations of the cohorts from which the samples were collected. (E) Bar plot for LARGE-PD Phase 2 with the proportions of AFR, EUR, EAS, SAS, and NAT ancestry clusters. Site names are listed in the Supplementary Information, Section Cohort Description. Cohort sample sizes are provided in Supplementary Table S1.

### Genotype-phenotype association study

#### Genome-wide association studies

Using SAIGE, three variants reached statistical significance on chromosome (chr) 4 in LARGE-PD P1, which replicated in P2 (Figure S13 (A), SI Table S4), with rs356183-G (inside the *SNCA* gene) emerging as the top SNP (P1 OR: 1.642, SE: 0.090, p-value: 3.45 × 10^−8^; P2 OR: 1.452, SE: 0.058, p-value: 1.27 × 10^−10^, Figure S14).

For ATT, we identified statistically significant associations in chr 3, 4, 11, and 13 in P1 (SI Figure S13(B), SI Table S5). However, only the chr 4 signals, which were observed in two different LD blocks (SI Figure S15), were replicated in P2. The top SNP in the first LD block was rs356182-G, the same variant reported on Loesch et al. (P1 OR: 1.736, SE: 0.082, p-value: 8.53 × 10^−12^, P2 OR: 1.467, SE: 0.057, p-value: 8.29 × 10^−12^). In the second block, the top SNP was rs6830166-C (P1 OR: 1.793, SE: 0.089, p-value: 2.06 × 10^−11^, P2 OR: 1.280, SE: 0.060, p-value: 3.14 × 10^−5^). Both SNPs are intronic variants inside the *SNCA* gene. Among the other regions, nine variants near the *NRROS* (chr 3) gene achieved statistical significance on P1, but exhibited an opposite effect in P2. Signals detected in other chr were not replicated.

For TRACTOR, we observed signals on *NRROS* and *SNCA* genes associated only with NAT ancestry (Figure S16(D), SI Table S6). Chr 4 exhibited two LD blocks (Figure S17), one led by rs3857049-T (P1 OR: 2.075, SE: 0.124, p-value: 1.70 × 10^−9^; P2 OR: 1.390, p-value: 7.52 × 10^−6^) and the other led by rs2737021-T (P1 OR: 0.484,SE: 0.128 , p-value: 7.75 × 10^−9^; P2 OR:0.632, SE: 0.088, p-value: 8.60 × 10^−8^). No variants on chr 3 were replicated.

### Meta-analysis

Random-effects meta-analyses were performed separately using SAIGE, ATT, and TRACTOR-NAT results (Table S7-S12). Across these methods, two genomic risk loci were consistently identified, mapped to *ITPKB* and *SNCA* genes (Figure 2).

**Figure 2:**
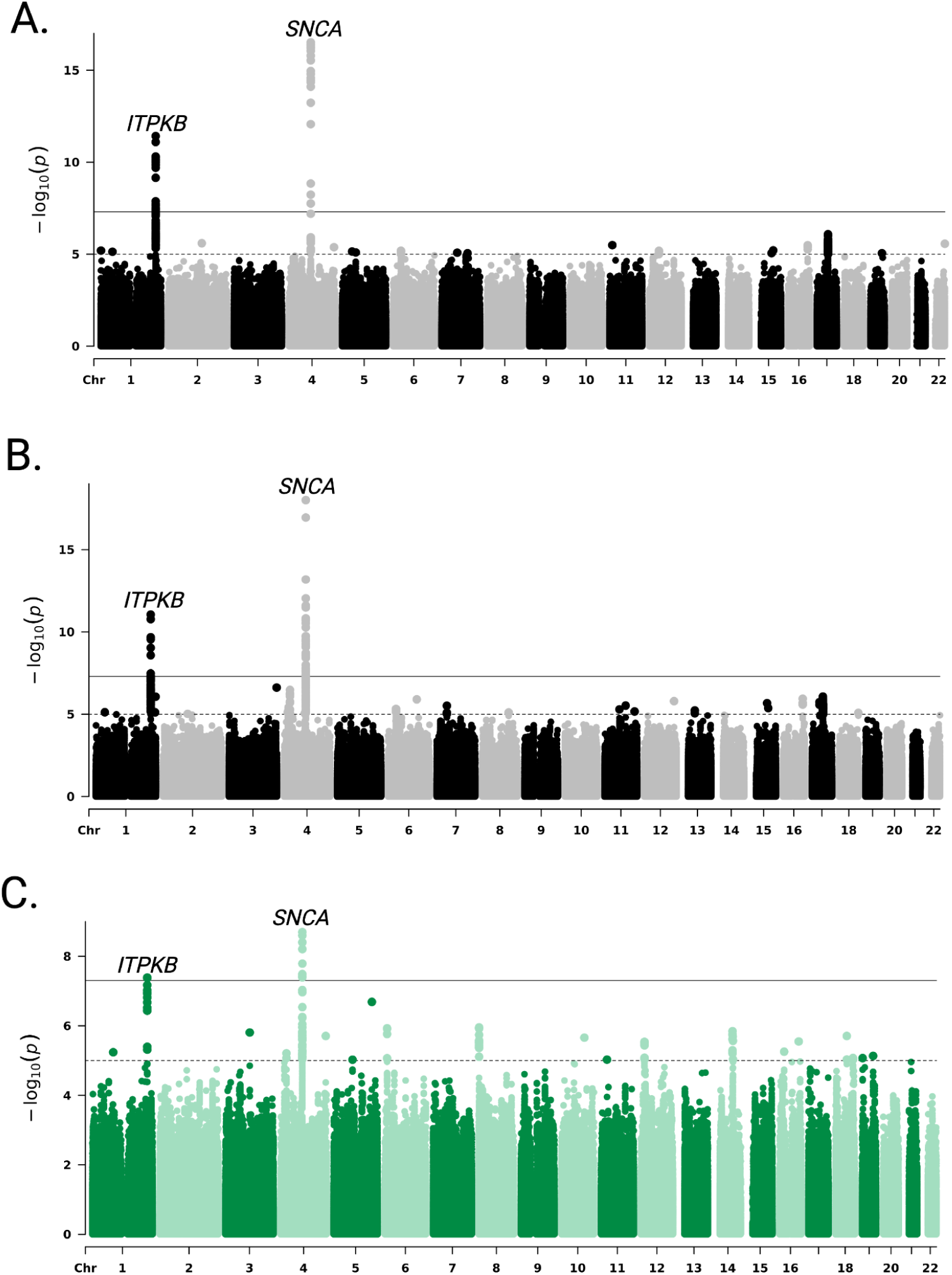
Manhattan plots from random-effects genome-wide association meta-analysis (GWAMA) of LARGE-PD Phase 1 and 2 cohorts for **(A**) SAIGE, **(B**) ATT, and (**C)** TRACTOR results for Native American ancestry meta-analysis. Each plot shows –log_10_(p) values across the genome; horizontal lines indicate genome-wide suggestive (1e-5) and statistical significance (5e-8) thresholds.

The SAIGE meta-analysis identified rs117185933-A as the top SNP on chr 1 (OR: 1.75, SE: 0.131, p-value: 3.80 × 10^−12^). On chr 4, two top SNPs were detected on *SNCA* gene: rs10027842-C (OR: 0.73, SE: 0.039, p-value: 1.76 × 10^−8^) and rs356223-G (OR: 0.664, SE: 0.031, p-value: 3.05 × 10^−17^).

The ATT meta-analysis identified rs16846413-T (OR: 1.679, SE: 0.118, p-value: 8.81 × 10^−12^) as the top variant on chr 1, and rs10027842-C (OR: 0.754, SE: 0.037, p-value: 4.17 × 10^−8^), also detected by SAIGE, along with rs356211-T (OR: 0.663, SE: 0.029, p-value: 9.83 × 10^−19^) on chr 4.

In the TRACTOR-NAT meta-analysis, the top variants were rs4653463-T (OR: 1.60, SE: 0.127, p-value: 4.09 × 10^−8^) on chr 1 and rs6532202-C (OR: 0.661, SE: 0.103, p-value: 1.62 × 10^−8^) and rs356181-A (OR: 0.6, SE: 0.047, p-value: 2.01 × 10^−9^) on chr 4. No variants reached statistical significance in the TRACTOR for EUR or AFR ancestry-specific meta-analyses (Figure S18).

The *ITPKB* loci (rs117185933-A) was previously associated with PD, being in LD with the lead SNP (rs74990530-T) in a recent SAS GWAS (figure S21). However, in our dataset both variants are independent lead signals (being s117185933-A the top hit in SAIGE meta-analysis) (table S17). Likewise, our meta-analysis with meta-regression of SAS and AMR cohorts indicate that rs117185933-A association is lost within random-effects meta-analysis, and is the lead MR-MEGA signal, with heterogeneity significantly correlated with ancestry (table S18). Furthermore, Bayesian fine mapping implemented by MR-MEGA nominates rs117185933-A as a putative causal variant being the single member of the 99% credible set for this genomic region (table S19), likely mapping an independent PD risk locus.

### Replication of previously identified PD risk loci

We investigated the beta coefficients and p-values for 78 and 90 GWAS hits reported by KIM and NALLS, respectively^7,8^. Among these, we identified 57 and 56 variants from the SAIGE (KIM: n=27; NALLS: n=30) and ATT (KIM: n=28; NALLS: n=28) meta-analyses, respectively, that had p < 0.05 and the same effect direction. One variant from KIM, rs79333841, had p < 0.05 but opposite effect directions in ATT and SAIGE (Figure S23). Variants with p > 0.05 were excluded from further consideration.

We also conducted a complementary analysis by selecting variants with p-values < 1 × 10^−5^ in our meta-analyses (SAIGE, ATT, and TRACTOR–NAT) and comparing their effect directions with variants showing p-values < 0.05 in KIM and/or NALLS. All overlapping variants from SAIGE (KIM n=13; NALLS: n=18), ATT (KIM: n=20; NALLS: n=36), and TRACTOR-NAT (KIM: n=3; NALLS: n=9) exhibited consistent effect directions (Figure S24).

### Admixture mapping

After performing AM, two significant regions were detected: one on chr 1, associated with EUR ancestry, and one on chr 6, associated with AFR ancestry (Figure S19). We then identified the genomic regions on chr1:242,089,864 - 243,560,064 and chr6:165,474,043 - 167,351,763. After performing the association study at the variant level, no variants on chr 1 achieved statistical significance, while 3 variants on chr 6 achieved statistical significance in P1, but were not replicated (SI Table S13).

### Functional annotation analysis

After using the MAGMA function available in FUMA, 16,992 gene ontology sets in MSigDB v7.0 were tested for enrichment of SNP signals based on the summary statistics provided. The ATT GWAMA gene-set analysis identified two significantly enriched gene-sets, belonging to the collections of Curated Gene Sets (Reactome Interleukin 23 signaling; BETA: 1.141, p-value: 1.463e-6, p-bon: 0.0248) and Gene Ontology Biological Process (Growth Hormone Receptor Signaling Pathway Via JAK-STAT; BETA 1.35, p-value 2.334e-6, p-bon: 0.0396).

After the replication and meta-analysis, we selected 15 variants identified as top or independent signals by FUMA (Table 2). For these variants, we retrieved CADD Phred scores, FORGEDB score, and SnpEff annotations. Most variants showed low CADD Phred scores and were annotated as intronic or intergenic according to SnpEff. However, the variant rs117185933 had a CADD Phred score of 17.92 (top ∼3% most deleterious variants) and FORGEdb 8, and was classified as a potential “5’ UTR premature start codon gain variant” according SnpEff (Table S14). Located 80 nucleotides upstream of the coding sequence in the 5′ untranslated region (UTR), this variant may introduce a premature start codon, potentially altering the translation initiation site. Such a change could result in a truncated or non-functional protein.

### Gene expression studies

We queried the rsIDs for the top or independent signals (Table 2) in the GTEx portal. No significant eQTL associations were identified for variants on chr 1. In contrast, eight variants on chr 4 showed significant associations with gene expression in multiple tissues. These included associations with the protein-coding genes *SNCA, SNCA-AS1*, and *MMRN1*, besides some long non-coding RNAs (Table S15).

Notably, eQTL associations were identified in several brain regions, including the amygdala, anterior cingulate cortex (BA24), caudate (basal ganglia), cerebellar hemisphere, frontal cortex (BA9), hippocampus, putamen (basal ganglia), and nucleus accumbens (basal ganglia). All regions showed associations with the *MMRN1* gene, with additional associations observed for *SNCA-AS1* in the anterior cingulate cortex and *SNCA* in the cerebellar hemisphere.

Associations were also observed in non-brain tissues, such as adipose tissue (subcutaneous and visceral - omentum), arteries (aorta and tibial), EBV-transformed lymphocytes, esophagus (mucosa), heart (atrial appendage and left ventricle), liver, skeletal muscle, tibial nerve, skin (not sun-exposed - suprapubic and sun-exposed - lower leg), and whole blood.

## Discussion

In this work we conducted the largest genotype-phenotype association study in Latin American populations. Our dataset is composed of data from two phases of LARGE-PD, the P1 with ∼1,500 samples and P2 with ∼4400. We performed GWAS with three different methods: i) SAIGE, which includes the GRM on the logistic regression; ii) ATT, which incorporates the GA on the statistical model; and iii) TRACTOR, which splits the dosage by ancestry based on LA information. Besides that, we also performed AM.

In our discovery cohort, we identified association signals on chr 3 (ATT and TRACTOR-NAT) and 4 (SAIGE, ATT, and TRACTOR-NAT); however, only the variants on chr 4 were successfully replicated. ATT and TRACTOR identified and replicated additional independent regions on chr 4 that SAIGE did not find. These findings show the importance of including ancestry information in our association models. Looking at previous studies, our effect directions were well correlated (r>0.69) with previous GWAS (Figure S23-S24), and most of our findings were also replicated in these prior studies (r>0.95).

The conducted meta-analysis using P1 and P2 data identified two genomic regions significantly associated with PD, located on chr 1 and 4. Notably, the region on chromosome 1 reached statistical significance in P2, but it did not achieve statistical significance in P1 (Table 2). Across meta-analyses, specific top SNPs and independent signals differed (SI Table S6-11). This likely reflects variations in analytical assumptions across the methods, while the consistent identification of *ITPKB* and *SNCA* underscores their robustness. Furthermore, random-effects meta-analysis found no evidence of between-cohort heterogeneity for the lead variants (I^2^ = 0%), except for TRACTOR-NAT on chromosome 4, with low-to-moderate I^2^ values (I^2^ = 11–37%) (Table 2). Because TRACTOR splits the genotypes by LA, differences in array content, LA inference, imputation methods, or gene-environment interactions may explain the observed heterogeneity.

Looking for the functional annotation in our study, we observed that the variant with the lowest p-value is inside the *SNCA* gene (Figure S20), which encodes Alpha-synuclein (α-Syn). These results are consistent with those of Loesch et al. GWAS^14^ and PRS^16^ findings, where rs365182 accounted for most of the trait variance in the PRS. α-Syn is a small protein (140 amino acids) involved in several key processes at the synapse, including modulation of the dopamine transporter ^45^, promotion of the dilation of the exocytotic fusion pore^45,46^, and others^47,48^. Notably, α-Syn has long been implicated in PD even before the advent of modern genetic studies^49,50^. In the GWAS era, the association became more evident^7,8,51^, with in vitro and in vivo models^52,53^ contributing to a deeper understanding of the role of α-Syn in PD. In PD cases, α-Syn accumulates in the substantia nigra, leading to the loss of dopaminergic neurons and the formation of Lewy bodies^54^. Interestingly, the *SNCA* gene was not associated with PD in the African GWAS^12^, highlighting the potential impact of genetic ancestry on PD genetic architecture.

We also identified variants in chr 1 within the *ITPKB* gene, which encodes the 3-kinase B of inositol 1,4,5-triphosphate (IP3). It influences key cellular processes, including calcium dynamics and immune function, and has been associated with neurodegenerative diseases, including Alzheimer’s disease^55^ and PD^55–58^. Some studies suggest that *ITPKB* may exert a protective effect against α-Syn aggregation^59^, and its expression levels are positively correlated with those of α-Syn^60^. Among the variants identified inside the *ITPKB* gene, rs117185933-A stands out with a high CADD Phred score. The functional annotation using SnEff classified this variant as a potential *5’ UTR premature start codon gain variant*. This variant shows its highest allele frequency in admixed American populations (8.35%), according to the gnomAD browser ^61^. The rs117185933-A was previously associated with PD, being in LD with the lead SNP (rs74990530).

Given that rs117185933-A was previously associated with PD, being in LD with the lead SNP (rs74990530-T) in a recent SAS PD GWAS^33^, and that both signals are independent in our SAIGE meta-analysis results (figure S21). A meta-analysis with meta-regression as implemented in Kim et al., 2024^8^ was performed to validate the independence of rs117185933-A across populations. Using both LARGE-PD cohorts and the two available SAS PD GWASs^33,34^, a random-effects meta-analysis and MR-MEGA were employed to uncover homogeneous and heterogeneous effects respectively. The rs117185933-A signal lost genome-wide significance under the random-effects model, showing high heterogeneity across cohorts that conditioned wider confidence intervals (table S18). However, MR-MEGA rescued rs117185933-A genome-wide significance while highlighting signal heterogeneity correlated with ancestry (p<0.05) and not residual heterogeneity (table S18), suggesting differences in population genetic structure drive the heterogeneity. Since MR-MEGA results identified rs117185933-A as the lead signal among other six independent SNPs (table S17), we leveraged the Bayes Factors provided to assemble credible sets as described previously^8^. Subsequently, rs117185933-A was identified as the single member of the 99% credible set, emerging as a novel putative causal variant that was not previously reported in the most recent multi-ancestry PD meta-analysis^8^.

Besides the single variant testing, gene-set-based MAGMA analysis through FUMA, based on ATT fixed and random-effects meta-analysis, identified two novel pathways associated with PD risk. Reactome Interleukin 23 signaling and Growth Hormone Receptor Signaling Pathway Via JAK-STAT were not previously reported among significant gene-sets in the most recent multi-ancestry meta-analysis for PD ^8^. However, a signal near the JAK1 gene was reported to be close to genome-wide significance under their meta-analysis ^8^. The biological role of JAK/STAT signaling has been highlighted in microglial activation and expression of pro-inflammatory cytokines, with therapeutic potential for its inhibition to halt neurodegeneration in PD ^62^. Hence, variants within members of JAK/STAT signaling-related pathways appear relevant for PD risk in LAC populations. Further studies to unveil their genomic significance are needed.

Our study has limitations. The sample size combined with the high genetic heterogeneity of Latin American populations, reduce statistical power to detect associations, especially for variants with small effect sizes. Additionally, the case-control imbalance in P2 (64% cases overall, 76% among males) may introduce bias. The cohorts include a high proportion of individuals from Peru (48% in P1 and 30% in P2). This uneven geographic representation may affect the generalizability of our findings to the broader LAC region, hence the relevance of continuing current efforts in the region. In terms of ancestry analysis, a major limitation is the underrepresentation of NAT individuals in reference panels. To compensate, we included non-admixed LARGE-PD controls, although this approach may introduce bias. Moreover, AFR ancestry is underrepresented in our dataset (5% in P1 and 2% in P2), reflecting a well-documented gap in PD research^63^. Regarding the meta-analysis with meta-regression and fine mapping approaches, variation in allele frequencies among SAS and LARGE-PD cohorts are taken as a proxy of ancestry to include in the meta-regression and different imputation panels were used by each study. Further validation with ancestry-matched LD estimates are needed, although the lack of robust panels for underrepresented and admixed populations will remain as a challenge. Likewise, MR-MEGA and random-effects meta-analysis of SAS and LARGE-PD cohorts outlined multiple independent signals besides the lead variants addressed. Hence, future follow up must assess the validity of rs117185933-A as a standalone PD putative causal variant within ITPKB and its biological mechanism.

Thus, our work contributes to elucidating the genetic architecture of PD in LAC populations by leveraging ancestry-based analyses. We successfully replicated findings from previous GWAS and identified novel variants associated with PD in this population, including rs117185933-A within the *ITPKB* gene. While these findings are promising, further investigation and validation are needed.

## Supporting information

Tables (main and SI)

## Data Availability

Data used in the preparation of this article were obtained from the Global Parkinson's Genetics Program (GP2; https://gp2.org) and from the Latin American Research Consortium on the Genetics of Parkinson's Disease (LARGE-PD) project. Specifically, we used Tier 2 data from GP2 release 10 (DOI 10.5281/zenodo.15748014, release 10). Tier 1 data can be accessed by completing a form on the Accelerating Medicines Partnership in Parkinson's Disease (AMP-PD) website (https://amp-pd.org/register-for-amp-pd). Tier 2 data access requires approval and a Data Use Agreement signed by your institution.

## Acknowledgments

This work was supported by the National Institutes of Health (NIH) grants (R01 1R01NS112499-01A1 to T.P.L, M.I-M, A.A., J.R., M.C-O and I.F.M.), Veterans Affair Puget Sound Healthcare System (5I01ABX005978-2 to T.P.L, V.B.P. and I.F.M.), ASAP-GP2 (M.C-O and I.F.M.), Michael J. Fox Foundation (MJFF-026283 for E.W., M.C-O and I.F.M.), Alzheimer’s Disease Sequencing Project (ADSP) (5U01AG076482-03 to E.W. and M.I-M), Vicedecanatura de Investigación y Extensión, Facultad de Medicina, Universidad Nacional de Colombia (for F.D.Z.), American Parkinson Disease Association (APDA) (1282087 for M.I-M and I.F.M; K.N.), Parkinson’s Foundation (PDGENE-1333334 M.I-M. and I.F.M; K.N.), National Council for Scientific and Technological Development (CNPq Brazil) (for B.L.S-L, STP, AFS, PBN and F.T.), DIEB-Universidad Nacional de Colombia (for G.A.), FAP-UNIFESP (for V.B.), Universidad Nacional de Tucumán (PIUNT D711 for C.A.), Vicerrectoría de Investigación, Universidad de Costa Rica (JFT), Centro de Investigaciones Clinicas (CIC) de la Fundación Valle del Lili (BMO), CONACYT-FORDECYT-PRONACES (11311 and 6390 for A.M-R), Programa de Apoyo a Proyectos de Investigación e Innovación Tecnológica–Universidad Nacional Autónoma de México (IN218023 for A.M-R and IN208622 for S.A.),Chan Zuckerberg Initiative Ancestry Network (2021-240438 for A.M-R), Secretaría de Ciencia, Humanidades, Tecnología e Innovación (Secihti) (CF-2019/6390. For S.A., and 1222481 for ALF), Margaret Q. Landenberger Foundation (for K.N.). This project was supported by the Global Parkinson’s Genetics Program (GP2). GP2 is funded by the Aligning Science Across Parkinson’s (ASAP) initiative and implemented by The Michael J. Fox Foundation for Parkinson’s Research (https://gp2.org). For a complete list of GP2 members see https://gp2.org.

The authors thank Emily Mason for her assistance with DNA extraction during the project, Dr. Daniel Shriner for his support on ancestry-related topics, and Sara Bandres Ciga with eQTLs and co-localization analysis. This work received technical support from Luis Aguilar, Alejandro León, and Jair García of the Laboratorio Nacional de Visualización Científica Avanzada at the Universidad Nacional Autónoma de México.

We also thank Carina Uribe Díaz and Alejandra Castillo Carbajal for their technical support at the Universidad Nacional Autónoma de México. We also acknowledge the technical and professional staff of the Biobank of Tissue and Fluids at the University of Chile for their commitment and support. We are grateful to the DNA-Neurogenetics Bank of the Instituto Nacional de Ciencias Neurológicas (INCN – Peru) for their support in the collection of DNA samples and associated data used in this publication. Finally, we are deeply grateful to all the individuals who participated in LARGE-PD.

## Data and code availability

Data used in the preparation of this article were obtained from the Global Parkinson’s Genetics Program (GP2; https://gp2.org) and from the Latin American Research Consortium on the Genetics of Parkinson’s Disease (LARGE-PD) project. Specifically, we used Tier 2 data from GP2 release 10 (DOI 10.5281/zenodo.15748014, release 10). Tier 1 data can be accessed by completing a form on the Accelerating Medicines Partnership in Parkinson’s Disease (AMP®-PD) website (https://amp-pd.org/register-for-amp-pd). Tier 2 data access requires approval and a Data Use Agreement signed by your institution.

All code generated for this article, and the identifiers for all software programs and packages used, are available on GitHub (https://github.com/GP2code/LARGE-PD_Phase2) and were given a persistent identifier via Zenodo (DOI: 10.5281/zenodo.15864760). Summary statistics from every ancestry-level meta-analysis are available on NDPK (https://ndkp.hugeamp.org/research.html?pageid=a2f_downloads_280).

## LARGE-PD consortium

Argentina: Emilia Mabel Gatto, Claudia Perandones, Martin Radrizani, Gustavo DaPrat, Natalia Gonzalez Rojas, Melisa Espindola, Martin Cesarini, Maria Valentina Muller, Carlos Matias López Razquin, Bibiana Pizarro, Lucia Wang, Clarisa Marchetti, Cesar Avila, Griselda Alvarado, Luciana Rojas-Vazquez, Juan Pablo Diaz-Rearte, Marcelo Kauffman, Sergio Rodriguez, Dolores Gonzales, Pavel Alejandro Hernandez, Belen Ceballos, Florencia Echeverria, Gabriela Costa, Marcelo Merello, Federico Capparelli, Florencia Wainberg, Tomas Poklepovich, Florencia Mallou, Denise De Belder

Bolivia: Erick Gonzalez, Enrique Wagner, Robin Rodriguez, Alexander Quecana Janco

Brazil: Bruno Lopes Santos-Lobato, Gracivane Lopes Eufraseo, Juliana Duarte, Marcella Montenegro, Tatiane Souza, Camille Sena, Ândrea Ribeiro-dos-Santos, Pedro Braga-Neto, Deborah Rangel, Marconny Cavalcante, Mateus Balsells, Vitor Tumas, Angela Vieira Pìmentel, Vanderci Borges, Carolina Candeias da Silva, Henrique Ballali Ferraz, Dayany Leonel Boone, Mariana Cavalcanti Costa, Egberto Reis Barbosa, Grace Letro, Artur F S Schuh, Carlos R M Rieder, Gabriela Magalhães Pereira, Deise Cristine Friedrich, Thalya Osmilda Alves de Carvalho, Isabella Fonseca Benati, Jullivan Käfer Pasin, Vitor Picanço Lima Gomes, Marcelo Somma Tessari, Ingrid Lorena da Silva Gomes, Juan Sebastián Sánchez León, Paula Saffie-Awad, Gabriel Alves Marconi, Manoella Guatimuzim Testa da Silva

Colombia: Gonzalo Arboleda, Oscar Bernal Pacheco, Oscar Bernal Pacheco, Tatiana Lopez Gonzalez, Humberto Arboleda, Carlos Eduardo Arboleda Bustos, Hebert Bernal Castro, Juan David Caicedo Narvaez, Kelly Bonilla Vargas, Jorge Orozco, Beatriz Munoz Ospina, Harold Londono, David Aguillon, Sonia Moreno, Omar Buritica, David Pineda, Marlene Jimenez-Del-Rio, Carlos Vélez-Pardo, Sarita Firstman

Chile: Pedro Chana-Cuevas, Ximena Pizarro Correa, Consuelo Moos, Natalia Rojas, Patricio Olguin, Alicia Colombo, Juan Cristobal Nuñez, Andres De la Cerda, María Francisca Canals, Gonzalo Farías, Valentina Bessa, Mérida Terán, Pen Cheng Zhongxomg, Paula Saffie, Eduardo Perez, Dominga Berrios, Elías Fernandez, Marlene Valenzuela Valenzuela, Mario Fuentealba Sandoval, Susana Pineda, Floria Pancetti, Maria Eugenia Contreras Pinto, Benjamin Soto Flores,

Costa Rica: Gabriel Torrealba-Acosta, Tanya Lobo-Prada, Jaime Fornaguera-Trías, Álvaro Hernandez-Guillen, Roger Rodríguez

Dominican Republic: Rossy Cruz Vicioso, Ernestina Castro, Alpher Perez, Sergio Mosquera, Cesarina Torres, Janfreisy Carbonell Honduras: Reyna M. Durón, Alex Medina, Heike Hesse, Evelin Alvarez Herrera, Eduardo P. Murillo, Glenda Oliva

El Salvador: Susana Pena, Tatiana Ascencio, Oscar Peña Rodas,

Ecuador: Faryd Llerena Toro, Michael Castelo, Carlos Rodriguez

Mexico: Mayela Rodríguez-Violante, Ana Jimena Hernández-Medrano, Amin Cervantes-Arriaga, Daniel Martinez Ramirez, Sarael Alcauter, Alejandra Medina-Rivera, Alejandra E. Ruiz-Contreras, Miguel E. Rentería, Alejandra Lázaro-Figueroa, Juan Manuel Esquivias-Farias, Andrés Morales-de-Arcia, Alejandra Zayas-Del Moral, Damaris Vazquez-Guevara, Yamil Matuk-Pérez , Carlos Manuel Guerra-Galicia, Ildefonso Rodriguez-Leyva, Karla Salinas-Barboza, Eugenia Morelos-Figaredo, Omar Cardenas-Saenz, Nadia A Gandarilla-Martínez, Sara Isais-Millán , Teresa Pérez-Torres, Domingo Martinez, Ingrid Estada-Bellmann, Roberto Trejo-Ayala, Carlos Alberto Ponce-Fernández, Dane Oropeza

Peru: Mario Cornejo-Olivas, Julia Rios Pinto, Angel Medina, Ivan Cornejo-Herrera, Koni Mejia-Rojas, Cintia Armas Puente, Edward Ochoa-Valle, Marcela Alvarado Morales, Elison Sarapura Castro, Andrea Rivera-Valdivia, Maryenela Illanes Manrique, Carla Manrique Enciso, Victoria Marca Ysabel, Olimpio Ortega Dávila, Freddy Requejo-Navarro, Alid Manrique Palomino, Gabriela Gushiken Oshiro, Laura Zelada Rios

Uruguay: Elena Dieguez, Victor Raggio, Andres Lescano

USA-Puerto Rico: Angel Vinuela, Esther Colon

USA: Thiago P Leal, Emily Waldo, Felipe Duarte-Zambrano, Miguel Inca-Martinez, Janvi Ramchandra, Kamilah Stark, Henry Mauricio Chaparro-Solano, Daniel Teixeira-dos-Santos, Emily Leininger, Nicolas Gutierrez, Valerie Rico, Paula Reyes-Pérez, Maria Rivera Paz, Ignacio F Mata, Karen Nuytemans, Anisley Martinez, Liena Infante

