## Supplementary material for "Genotype-phenotype association study conducted on LARGE-PD reveals novel loci associated with Parkinson’s Disease": SI text

### **Supplementary information of Genotype-Phenotype association studies in Latin American Cohort reveals new loci associated with Parkinson’s Disease**

#### Cohort description

The LARGE-PD Phase 2 study consists of seven genotyping batches from ten countries, totaling 23 sites (Table 1, Table S1-S3, Figure 1).

**LARGE-PD South America** is composed of samples from:

Argentina

- Hospital Sanatorio de la Trinidad Mitre (AR)
- Hospital Interzonal General de Agudos San Martin de La Plata (EZ)
- Hospital Angel C Padilla (TU)

Brazil

- Hospital Ophir Loyola (BE)
- Hospital e Maternidade Celso Pierro (HMCP) da Sociedade Campineira de Educação e Instrução (CA)
- Hospital Universitário Walter Cantídio (FT)
- Hospital de Clínicas de Porto Alegre (PO)
- Hospital de Clínicas de Ribeirão Preto (RP)
- Universidade Federal de São Paulo (SA)

Chile

- Centro de Trastornos del Movimiento (CH)
- Universidad de Chile (UC)

Colombia

- Universidad Nacional de Colombia (CB)
- Fundación Valle Del Lili (CC)
- Universidad de Antioquia (CO)

Peru

- The Peruvian cohort (PP) is composed of multiple institutions:
- Instituto Nacional de Ciencias Neurológicas
- Hospital Domingo Olavegoya, Jauja
- Dirección Regional de Salud, Puno
- Hospital Hipólito Unanue, Tacna
- Hospital Regional de Cusco, Cusco
- Centro Privado de Educación Medica Continua EDMECON

Uruguay

- Universidade de la República (UR)

**LARGE-PD Central America and Caribbean** is composed of samples from:

Costa Rica

- Universidad de Costa Rica (CR)

Honduras

- Hospital de Especialidades San Felipe (HO)

**LARGE-PD North America** is composed of samples from:

Mexico

- Instituto Nacional de Neurología y Neurocirugía Manuel Velasco Suárez (MI)
- Instituto Tecnológico y de Estudios Superiores de Monterrey (MX)
- Universidad Nacional Autónoma de México (UM)

United States of America

- Fundación Parkinson Puerto Rico (PR)
- University of Miami (US)

#### Genotyping calling and Quality Control

As mentioned in the main text, genotype calling was performed using the Illumina Array Analysis Platform (IAAAP) with the GP2 cluster file. We chose to use this cluster file instead of the one provided by Illumina because cluster files generated from larger studies tend to increase sample call rates and reduce the likelihood of miss-clustering ^1^.

As part of the genotyping calling QC, we removed unaligned variants (i.e., those mapped to chromosome 0) and other problematic variants. Problematic variants were defined as those that did not genotype the reference allele or that changed chromosomes when comparing different human genome builds. Variants lacking the reference allele,i.e., variants that genotyped only alternative alleles, were identified through comparison with the dbSNP database. LiftOver-related issues were detected by comparing two annotation tables for the NeuroBooster Array (NBA), one aligned to hg37 and the other to hg38.

#### Relationship control

We split the QCed dataset by collection site for each LARGE-PD dataset, calculated the genetic relationship matrix using KING ^2^, and generated the list of samples to be removed, excluding all second-degree or higher relationships (kinship cutoff > 0.0884) using NAToRA ^3^. All NAToRA analysis was performed using the optimal version, ie, it removed the smallest number of samples possible.

#### Ancestry Analysis

Latin American and Caribbean (LAC) populations are the result of admixture over the past ~500 years, primarily involving African, European, and Native American ancestries ^4–6^. This admixture introduces genetic heterogeneity, which can confound association studies. To address this, we performed ancestry analysis to better account for the population structure and demographic complexity.

In this section, we explain methodological details about the ancestry analysis performed in this paper. All analysis is based on a reference (1KGP 30x) and a target dataset (LARGE-PD Phase 1 or LARGE-PD Phase 2). The samples from 1KGP samples used as reference can be accessed on https://github.com/MataLabCCF/GnomixPretrained/blob/main/Parentals/.

The ancestry analysis is composed of three sections: (i) Principal Component Analysis, (ii) ADMIXTURE Analysis, and (iii) Local Ancestry Analysis

##### Principal Component Analysis

Principal Component Analysis (PCA) is a crucial component for genotype-phenotype association studies, as it allows for population structure control, thereby reducing the incidence of false positives and false negatives in the association results. PCA is a dimensionality reduction technique that transforms data into principal components representing the variation within the dataset.

To perform the projected Principal Component Analysis (PCA), we first generated a list of variants in common between the reference dataset (1KGP) and the target dataset. These datasets were then merged, and variants with minor allele frequency (MAF) < 0.01 or in linkage disequilibrium (LD) were removed using PLINK v2.00a5.10LM ^7^. The remaining variants after filtering were extracted from both the reference and target QCed data to be used in the projected PCA.

For the projected PCA computation using GCTA ^8^, we first calculated the genetic relationship matrix (GRM) using the reference data and subsequently performed PCA inference based on the GRM. Next, we calculated the principal component (PC) loadings for each variant, which were then used to compute the PCs on the target dataset. Using this pipeline, we inferred 50 PC for LARGE-PD Phase 1 and Phase 2.

The projected PCA was inferred separately for LARGE-PD Phase 1 and LARGE-PD Phase 2 because these datasets were genotyped using different arrays (MEGA for Phase 1 and NeuroBooster for Phase 2). This separation was necessary to address the reduced number of overlapping variants that could arise due to differences in genotyping platforms.

For PCA without projection, we removed variants with minor allele frequency (MAF) < 0.01 and variants in linkage disequilibrium (LD). After this, we calculated the genetic relationship matrix (GRM) using the reference data and subsequently performed PCA inference based on the GRM.

##### ADMIXTURE Analysis

The ADMIXTURE ^9^ is a methodology for inferring population structure by analyzing genetic data. It identifies clusters within a dataset of genetic samples, with two distinct modes of operation. In the **unsupervised mode**, clusters are inferred solely based on the target dataset, without any prior information about population structure. In contrast, the **supervised mode** requires a list of non-admixed samples to be used as reference. This reference dataset guides the clustering process on the target dataset.

For the supervised ADMIXTURE analysis, we first generated a list of variants in common between the reference and target datasets. These datasets were then merged, and variants in LD were removed. The processed dataset was exported in PLINK binary format (bed/bim/fam files). A population file was created, specifying the parental population assignment for each reference sample.

ADMIXTURE was executed in supervised mode using the --supervised flag using k=5 (same number of parental populations). The analysis was repeated 10 times, and the run with the lowest cross-validation error was selected to be analyzed.

Our analysis showed that our populations are typically three-way admixed, with varying AFR, EUR, and NAT ancestry proportions. In LARGE-PD, the ancestry composition of PD cases differs between phases, with P1 cases comprising 4.0% AFR, 44.9% EUR, and 49.9% NAT, while P2 cases have 5.4% AFR, 54.0% EUR, and 39.9% NAT. Similarly, the ancestry proportions among controls vary, with P1 controls showing 5.3% AFR, 50.6% EUR, and 43.6% NAT, whereas P2 controls exhibit 2.5% AFR, 28.5% EUR, and 68.5% NAT. EAS and SAS contributed to less than 1% of the total ancestry in both phases (Figure 1, SI Table S3).

##### Local Ancestry

The chromosomes of an admixed individual form a mosaic of fragments from different ancestries. These fragments, known as ancestry tracts ^10^, vary in size depending on the timing of the admixture event and the length of the chromosome.

To obtain the most reliable Local Ancestry (LA) inference, we started testing for the best possible panel to use as our reference dataset. Our first test compared the five-way (AFR (N=471), EUR (N=136), EAS (N=316), SAS (N=67) , NAT (N=23)) reference from 1KGP against the three-way reference panel from 1KGP (AFR, EUR, NAT).

We trained the model and compared the confusion matrices generated by G-Nomix for the five-way and three-way reference panels using two chromosomes (4 and 17) (Figure S25). Based on the ADMIXTURE results and the confusion matrices, we concluded that a 3-way admixed panel was the most suitable choice for our model. After selecting the 3-way admixed panel, we searched for non-admixed individuals to enhance the Native American (NAT) reference panel. We included LARGE-PD control samples identified as non-admixed based on ADMIXTURE and projected PCA analyses, resulting in the addition of 69 individuals from Phase 1 and 158 from Phase 2.

Subsequently, we tested variations in two parameters: model type ("best" or "default") and window size (0.3 cM, 0.5 cM, and 0.7 cM) (Figure S26). This analysis was performed across all chromosomes for all LARGE-PD Phase 2 samples. Posteriori values were compared across three chromosomal regions: the beginning (first third of total windows), the middle, and the end (last third of total windows). We ultimately chose the model "best" with a window size of 0.5 cM. The pretrained models and the list of samples are publicly available at https://github.com/MataLabCCF/GnomixPretrained.

#### Admixture Mapping

Our Admixture Mapping (AM) pipeline consists of converting the MSP file into a VCF-like format for each parental population, and the association analysis was performed using the GENESIS R package^11^. We conducted a joint test, where all ancestries were tested simultaneously in an AM logistic mixed model, as well as per-ancestry tests to identify which parental population was responsible for the association signal. Our null model included age, sex, 10 PCs, and the GRM inferred using KING ^12^.

The p-value threshold was determined based on the number of independent tests, using the same method applied in our previous X-Chromosome Wide Association Study ^13^. Specifically, we calculated the correlation matrix for all variants in our VCF-like file, estimated the number of independent tests, and applied the Bonferroni correction.

The pipeline is publicly available at <https://github.com/MataLabCCF/GenesisAM>.

#### Meta-analysis

To meta-analyze the GWAS results from LARGE-PD Phases 1 and 2, we implemented modifications to the pipeline used in previous studies ^13^. Changes were made to accommodate the format of our summary statistics and to account for the group of variants that have not been assigned an rsID by the TOPMed imputation server, which prevented the pipeline from running. Such variants were excluded from the SAIGE meta-analysis; none were statistically significant (p-value range 5e-4 - 0.9) and are described in detail in Table S15.

Additionally, no genomic control (GC) was applied because each GWAS included the first ten PCs as covariates, and this approach has proven more effective at accounting for population stratification ^14^.

#### Functional annotation

FUMA (Functional Mapping and Annotation) is a tool that aims to facilitate post-GWAS analysis. It uses a two-step clumping strategy to define independent and lead variants. Initially, independent signals are identified by excluding variants in high linkage disequilibrium (LD) (r² > 0.6). In a second step, a stricter LD pruning is applied, in which variants that are not in LD with each other (r² < 0.1) and located within a 250 kb window are considered lead variants^15^.

CADD is a tool that integrates multiple annotations to predict the deleteriousness of genetic variants. Since raw CADD scores are not directly interpretable, we used the Phred-scaled CADD scores (rank-based metric), which higher scores indicate a greater likelihood that a variant is deleterious. SnpEff is a tool that annotates genetic variants and predicts their effects on genes, such as amino acid changes. It also classifies the functional consequences by estimating their impact on gene function. SnpEff annotations were retrieved via the MyVariant.info platform ^16^.
