## Supplementary material for "Genotype-phenotype association study conducted on LARGE-PD reveals novel loci associated with Parkinson’s Disease": SI Figures

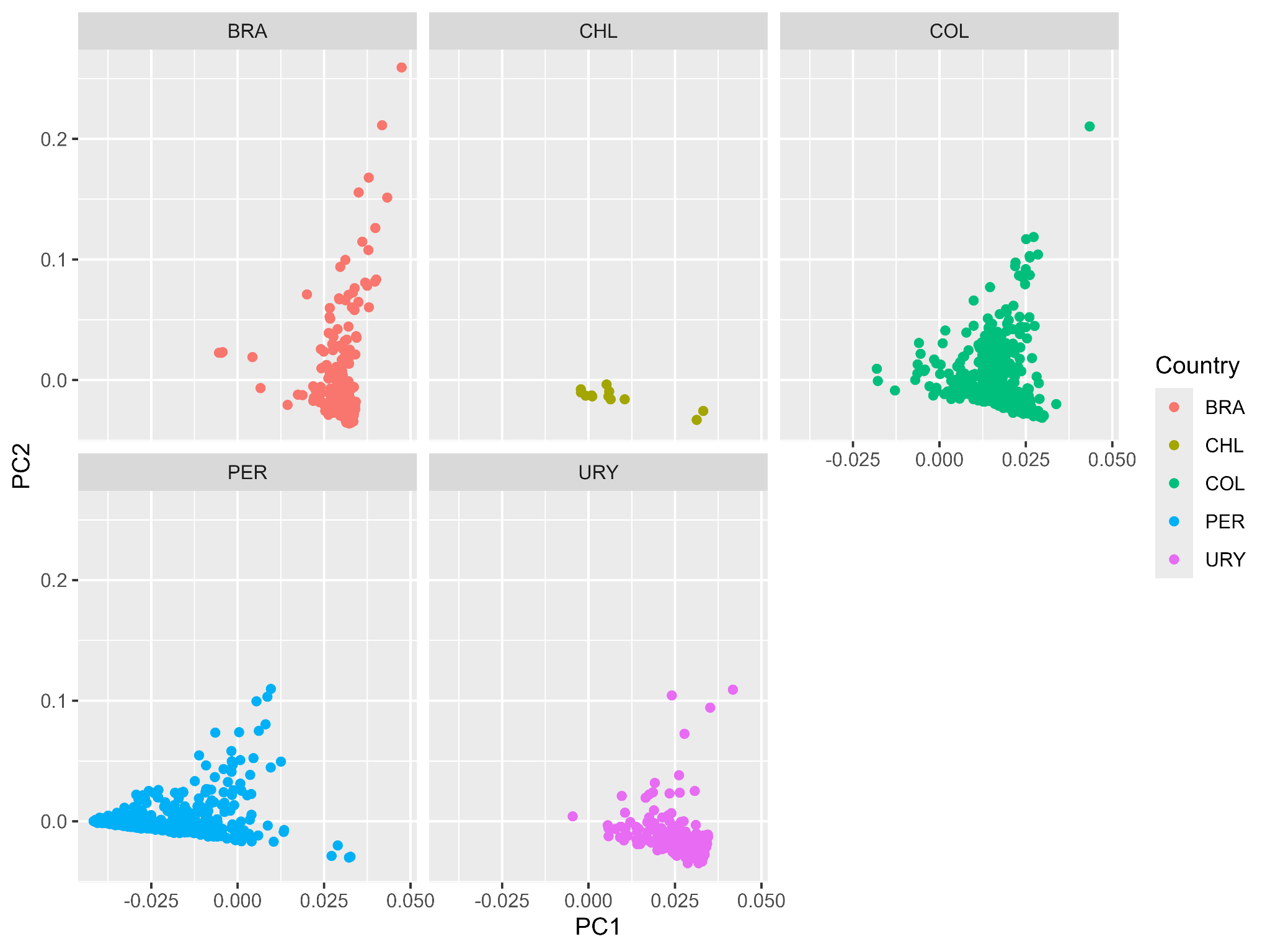

### **Figure S1:** Principal Component Analysis (PCA) for LARGE-PD Phase 1 (PC1 vs. PC2). The PCA was performed using all samples and is plotted by country for visual clarity. BRA: Brazil, CHL: Chile, COL: Colombia PER: Peru, PRI: Puerto Rico, URY: Uruguay.

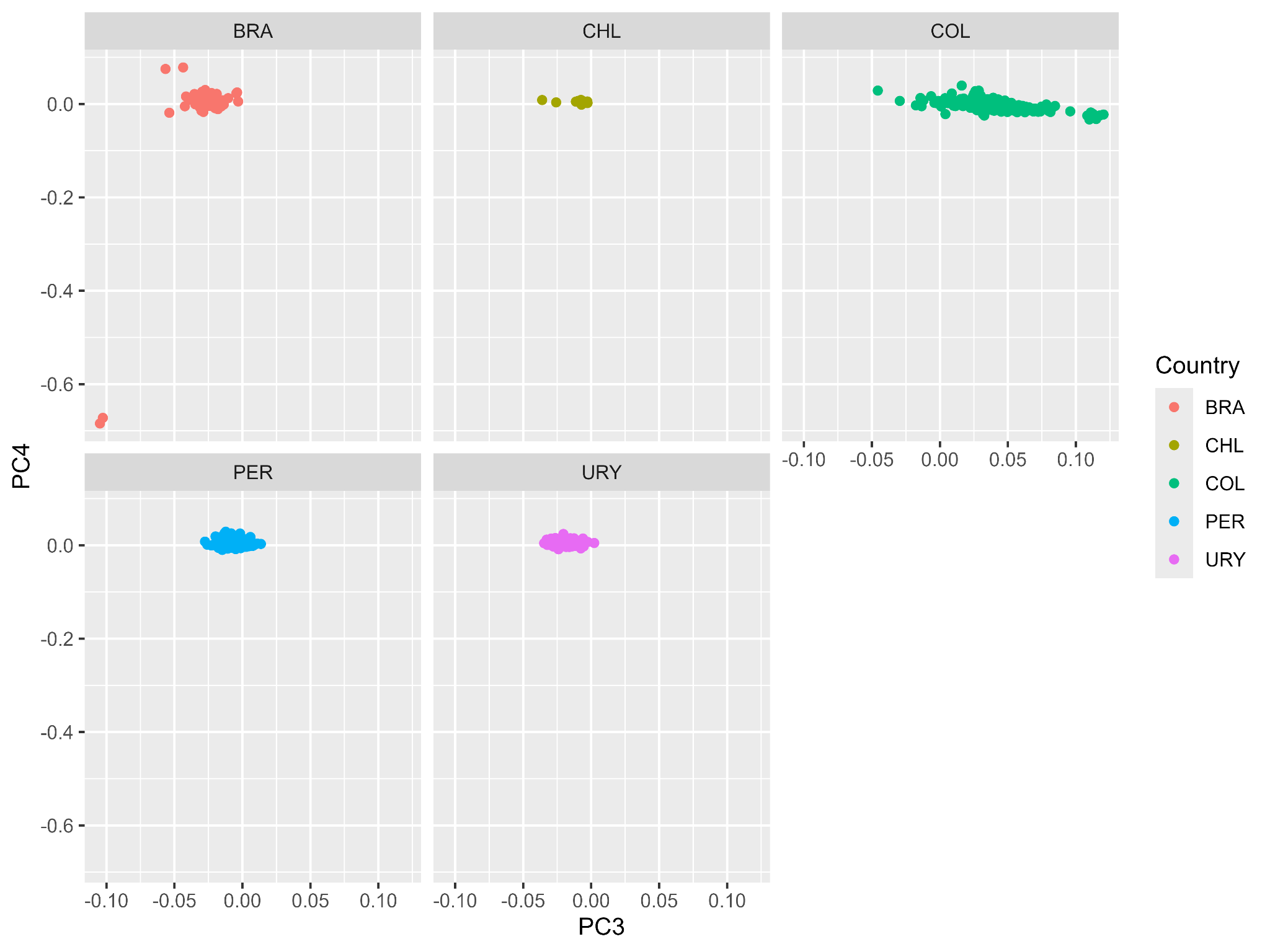

### **Figure S2:** Principal Component Analysis (PCA) for LARGE-PD Phase 1 (PC3 vs. PC4). The PCA was performed using all samples and is plotted by country for visual clarity. BRA: Brazil, CHL: Chile, COL: Colombia PER: Peru, PRI: Puerto Rico, URY: Uruguay.

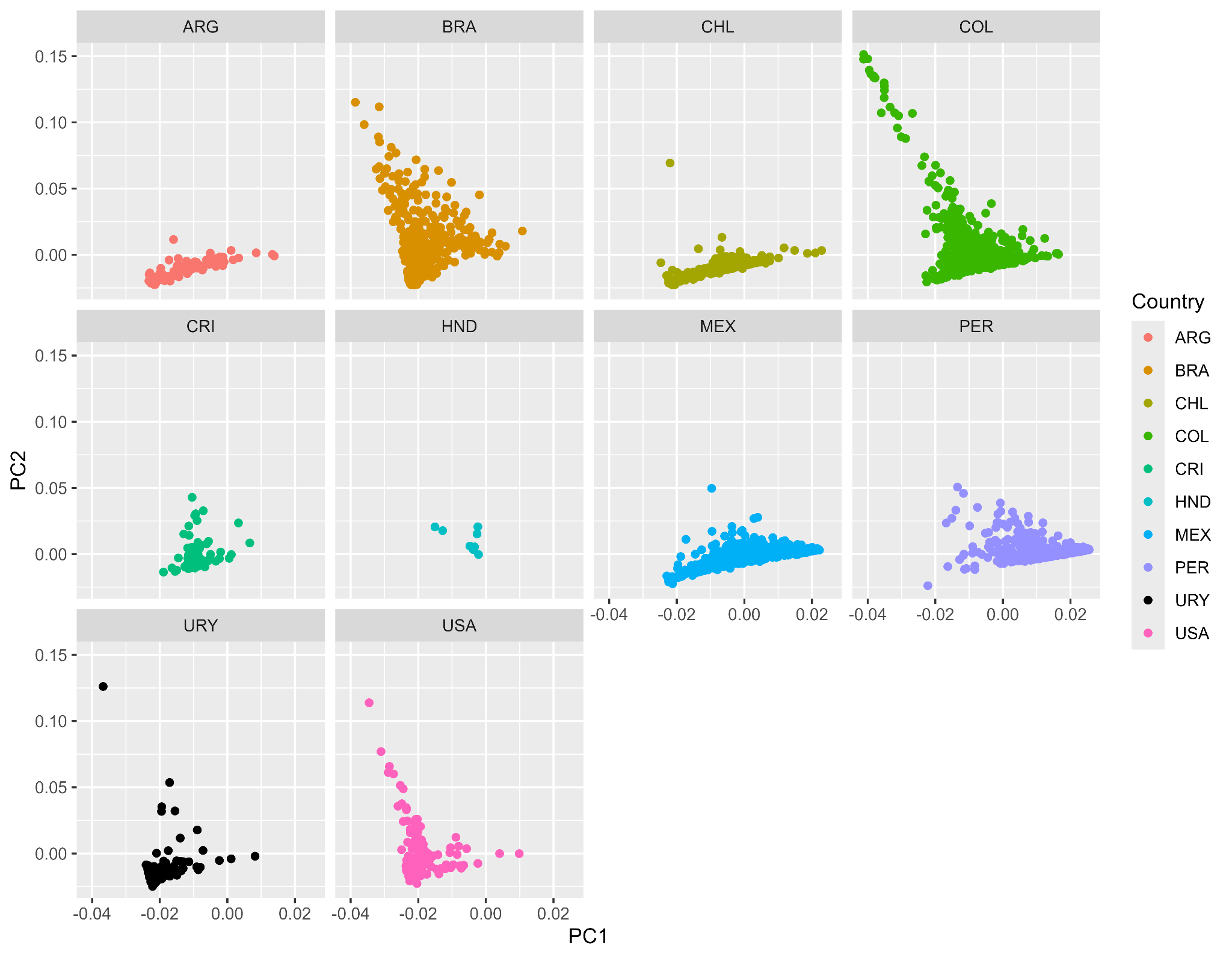

### **Figure S3:** Principal Component Analysis (PCA) for LARGE-PD Phase 2 (PC1 vs. PC2). The PCA was performed using all samples and is plotted by country for visual clarity. ARG: Argentina, BRA: Brazil, CHL: Chile, COL: Colombia, CRI: Costa Rica, HND: Honduras, MEX: Mexico, PER: Peru, URY: Uruguay, USA: United States of America

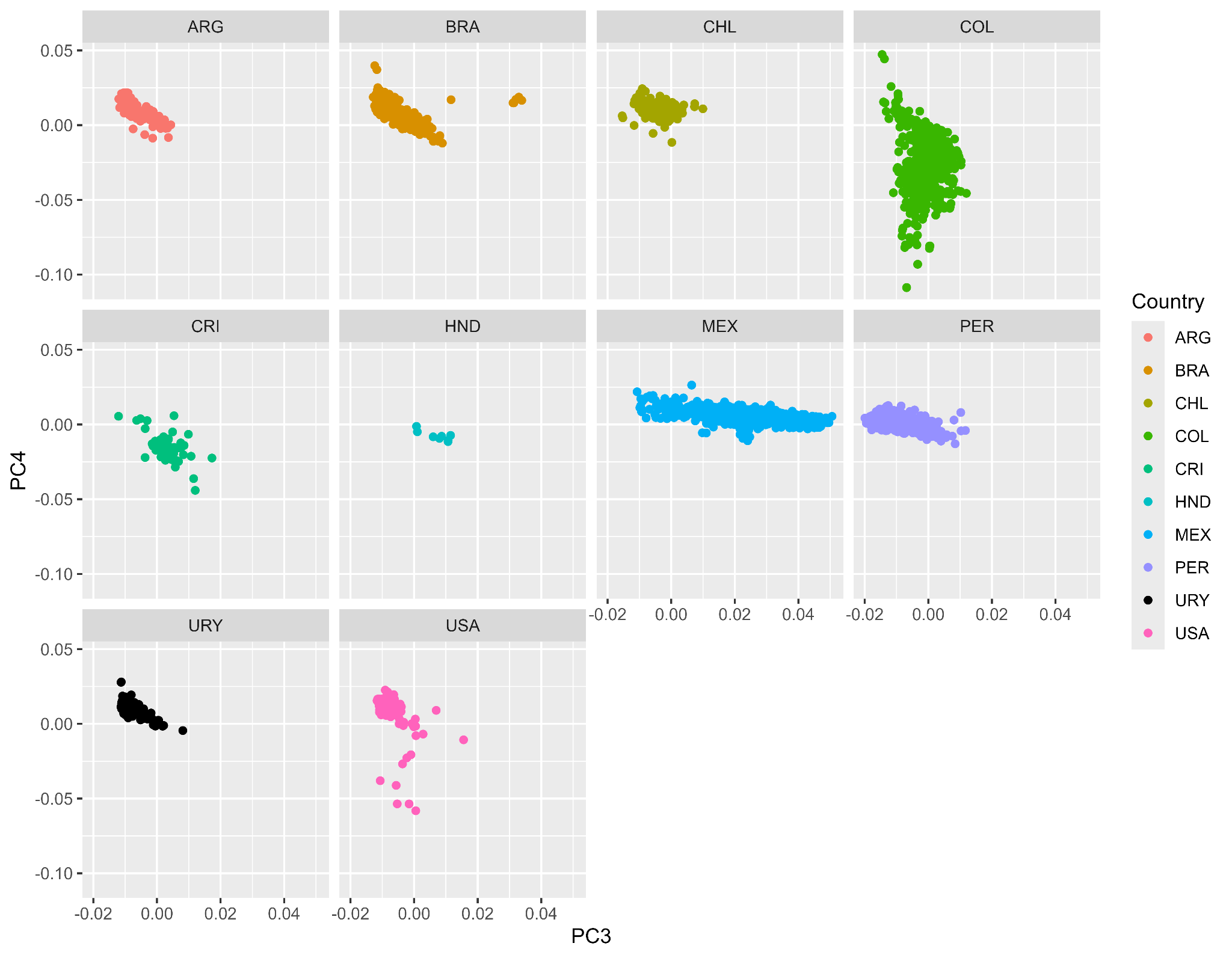

### **Figure S4:** Principal Component Analysis (PCA) for LARGE-PD Phase 2 (PC3 vs. PC4). The PCA was performed using all samples and is plotted by country for visual clarity. ARG: Argentina, BRA: Brazil, CHL: Chile, COL: Colombia, CRI: Costa Rica, HND: Honduras, MEX: Mexico, PER: Peru, URY: Uruguay, USA: United States of America

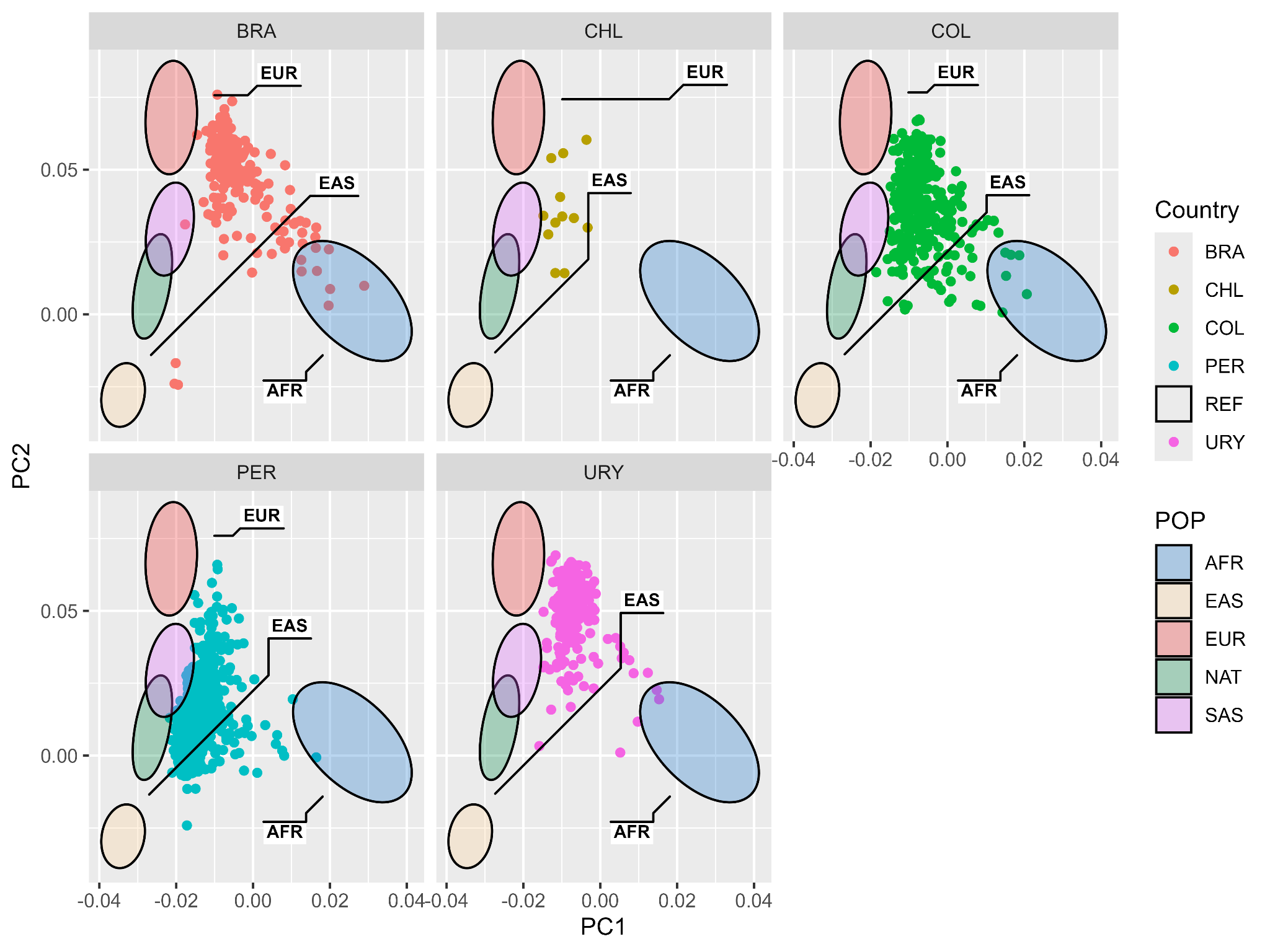

### **Figure S5:** Projected PCA for LARGE-PD Phase 1 (PC1 and PC2). The ellipses represent the parental population. The PCA was inferred with all samples and plotted per country for visual clarity. BRA: Brazil, CHL: Chile, COL: Colombia, PER: Peru, URY: Uruguay.

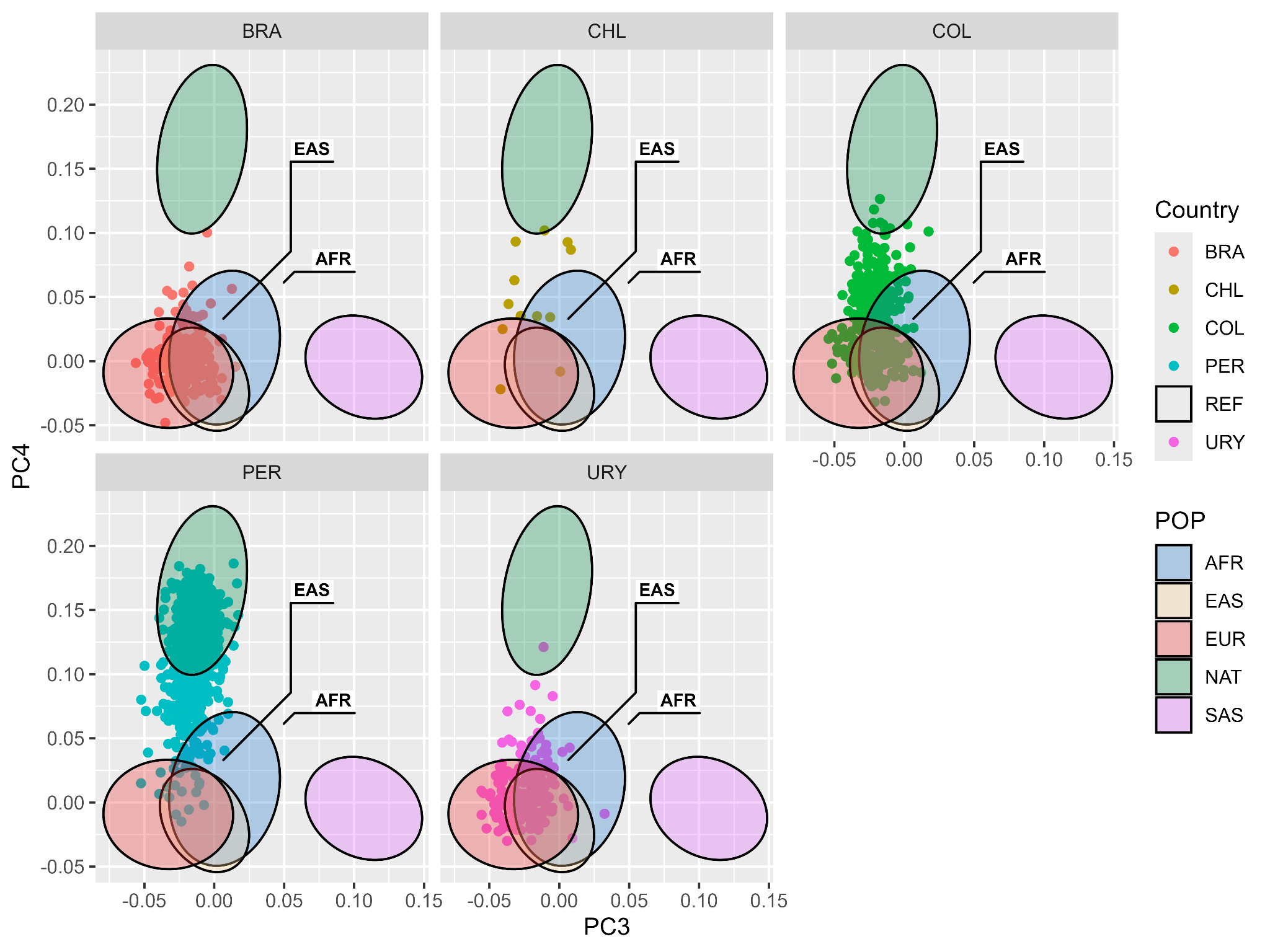

### **Figure S6:** Projected PCA for LARGE-PD Phase 1 (PC3 and PC4). The ellipses represent the parental population. The PCA was inferred with all samples and plotted per country for visual clarity. BRA: Brazil, CHL: Chile, COL: Colombia, PER: Peru, URY: Uruguay.

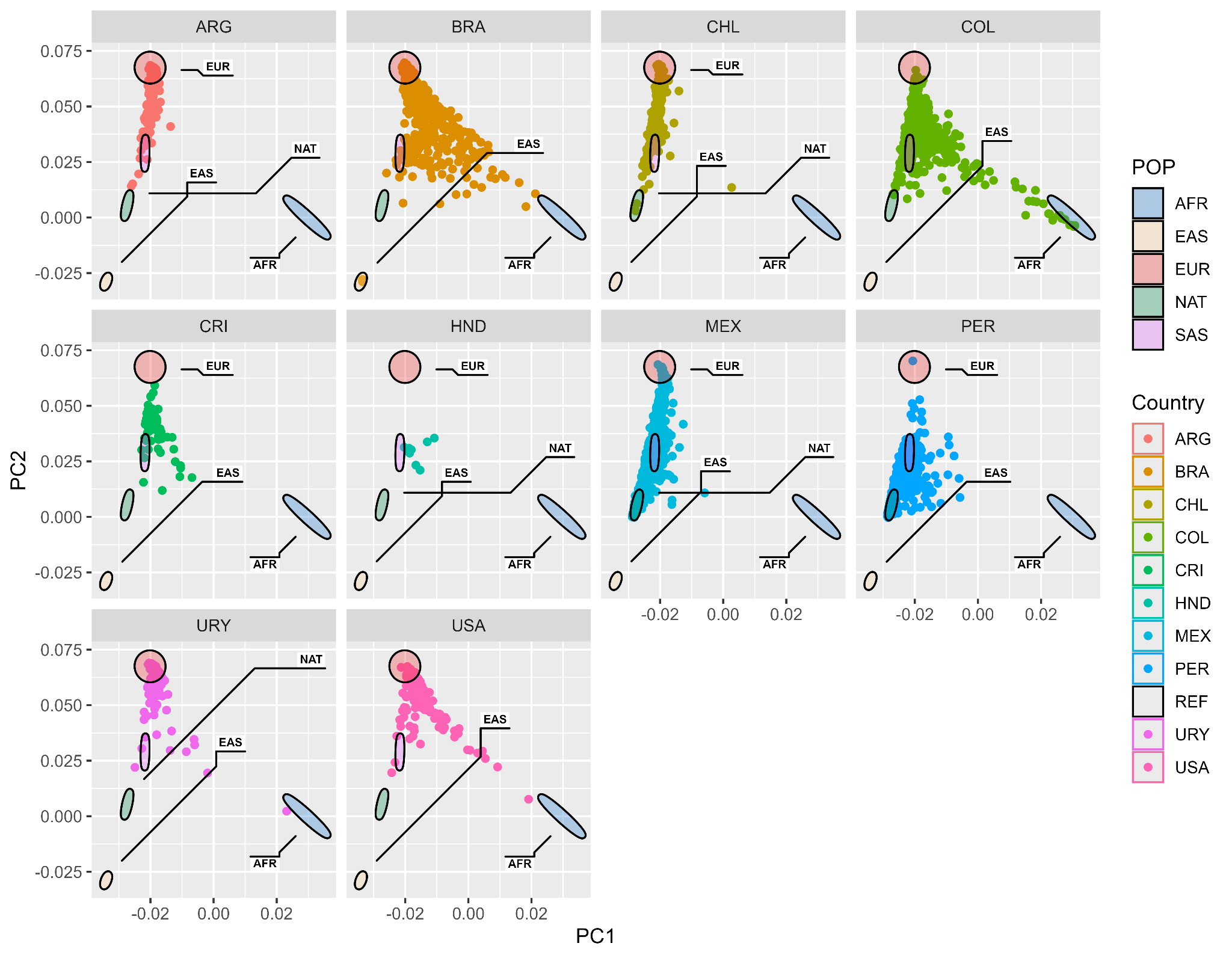

### **Figure S7:** Projected PCA for LARGE-PD Phase 2 (PC1 and PC2). The ellipses represent the parental population. The PCA was inferred with all samples and plotted per country for visual clarity. ARG: Argentina, BRA: Brazil, CHL: Chile, COL: Colombia, CRI: Costa Rica, HND: Honduras, MEX: Mexico, PER: Peru, URY: Uruguay, USA: United States of America

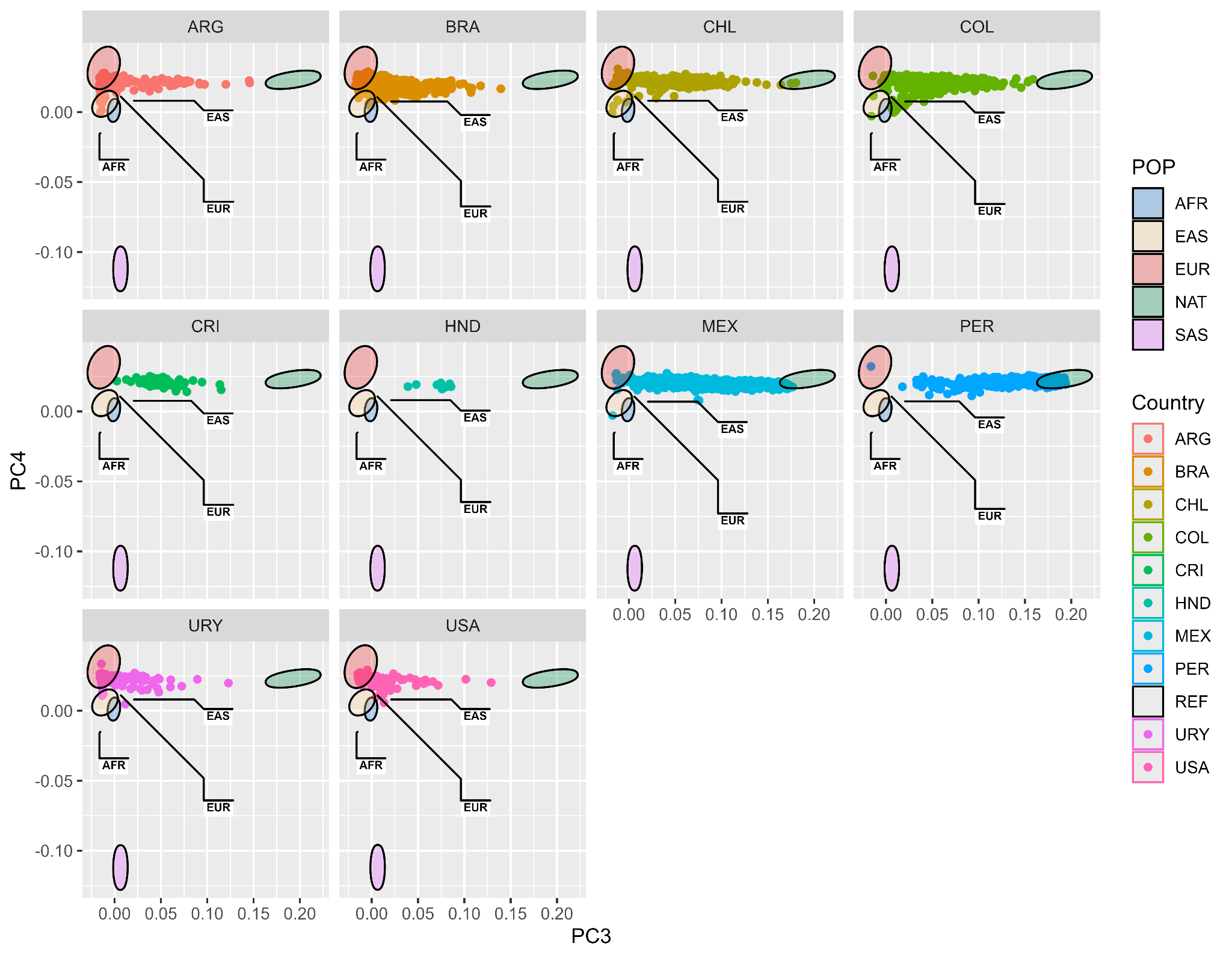
**Figure S8:** Projected PCA for LARGE-PD Phase 2 (PC3 and PC4). The ellipses represent the parental population. The PCA was inferred with all samples and plotted per country for visual clarity. ARG: Argentina, BRA: Brazil, CHL: Chile, COL: Colombia, CRI: Costa Rica, HND: Honduras, MEX: Mexico, PER: Peru, URY: Uruguay, USA: United States of America

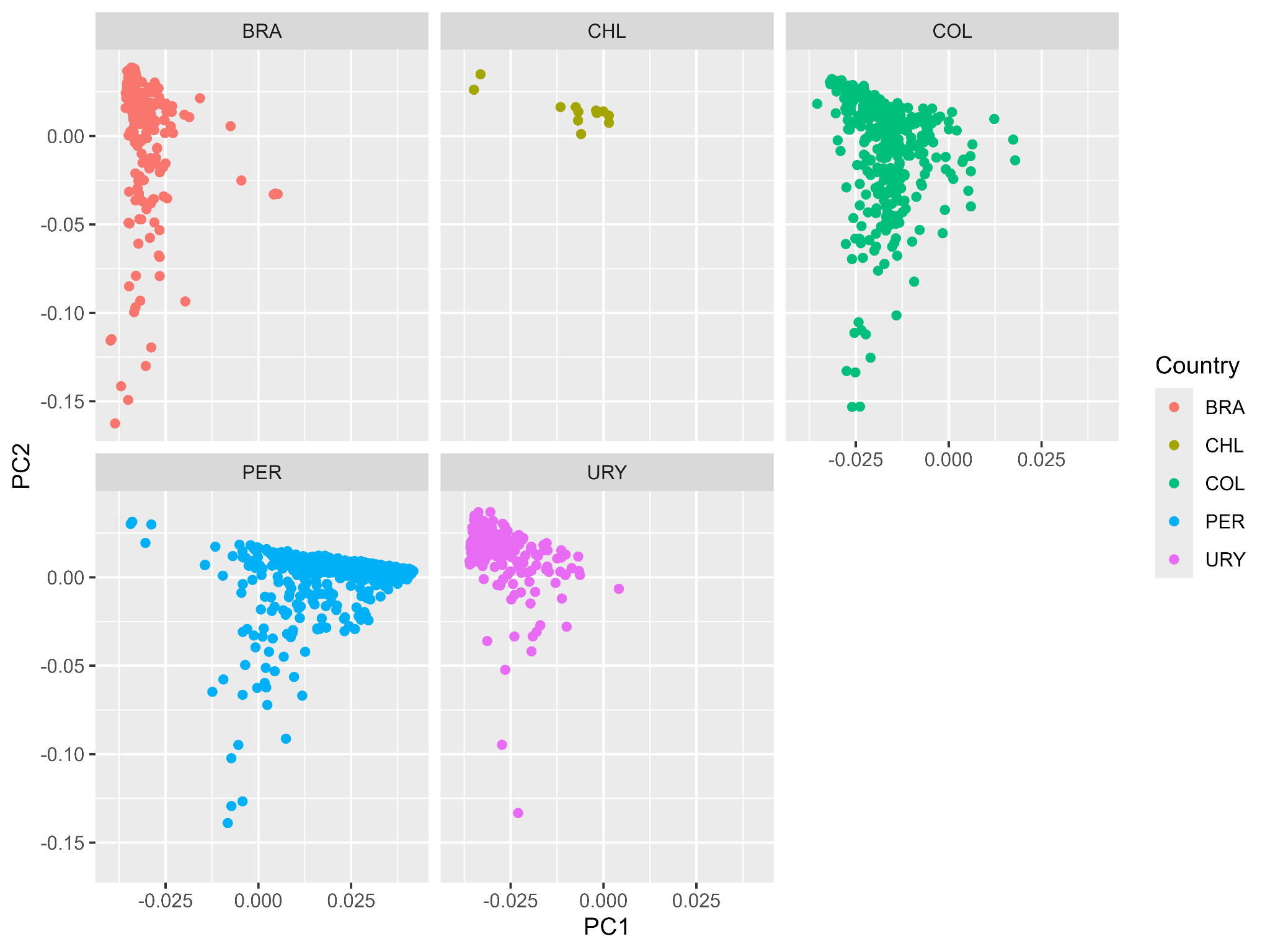

### **Figure S9:** Principal Component Analysis (PCA) for LARGE-PD Phase 1 (PC1 vs. PC2) excluding outliers detected across the first 10 principal components from the projected PCA of LARGE-PD Phase 1. The PCA was performed using all samples and is plotted by country for visual clarity. BRA: Brazil, CHL: Chile, COL: Colombia, PER: Peru, PRI: Puerto Rico, URY: Uruguay.

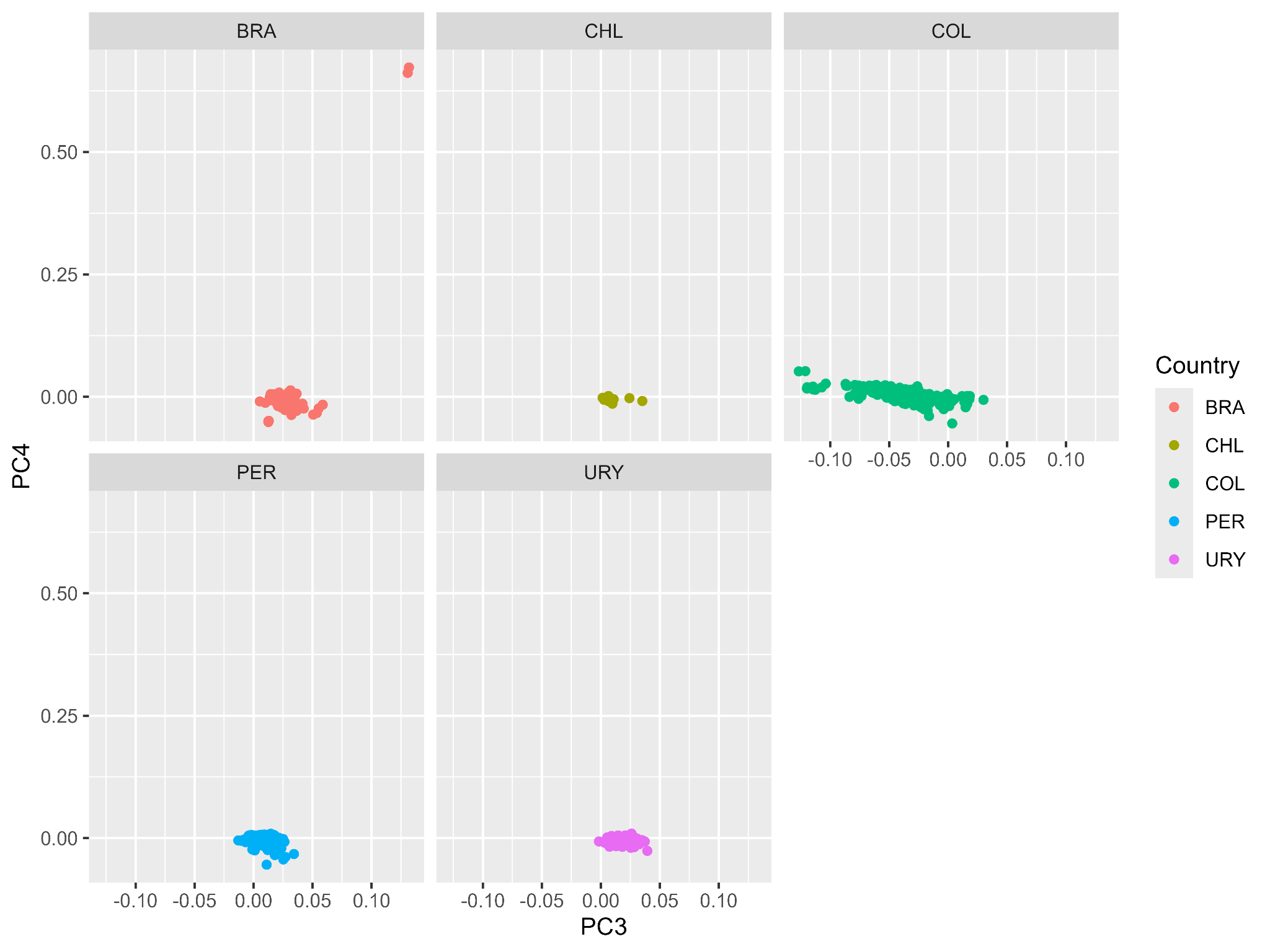

### **Figure S10:** Principal Component Analysis (PCA) for LARGE-PD Phase 1 (PC3 vs. PC4) excluding outliers detected across the first 10 principal components from the projected PCA of LARGE-PD Phase 1. The PCA was performed using all samples and is plotted by country for visual clarity. BRA: Brazil, CHL: Chile, COL: Colombia, PER: Peru, PRI: Puerto Rico, URY: Uruguay.

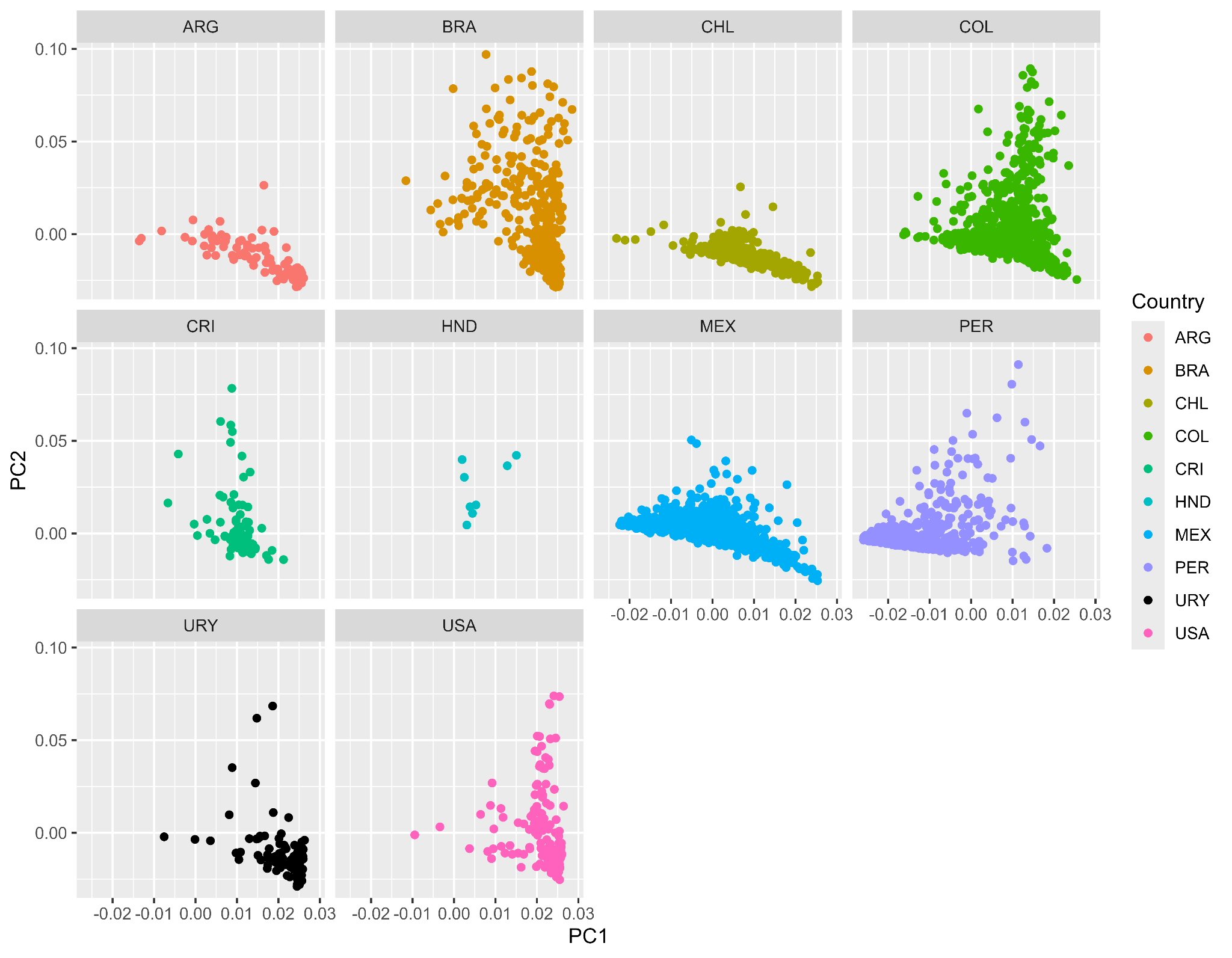

### **Figure S11:** Principal Component Analysis (PCA) for LARGE-PD Phase 2 (PC1 vs. PC2) excluding outliers detected across the first 10 principal components from the projected PCA of LARGE-PD Phase 2. The PCA was performed using all samples and is plotted by country for visual clarity. ARG: Argentina, BRA: Brazil, CHL: Chile, COL: Colombia, CRI: Costa Rica, HND: Honduras, MEX: Mexico, PER: Peru, URY: Uruguay, USA: United States of America

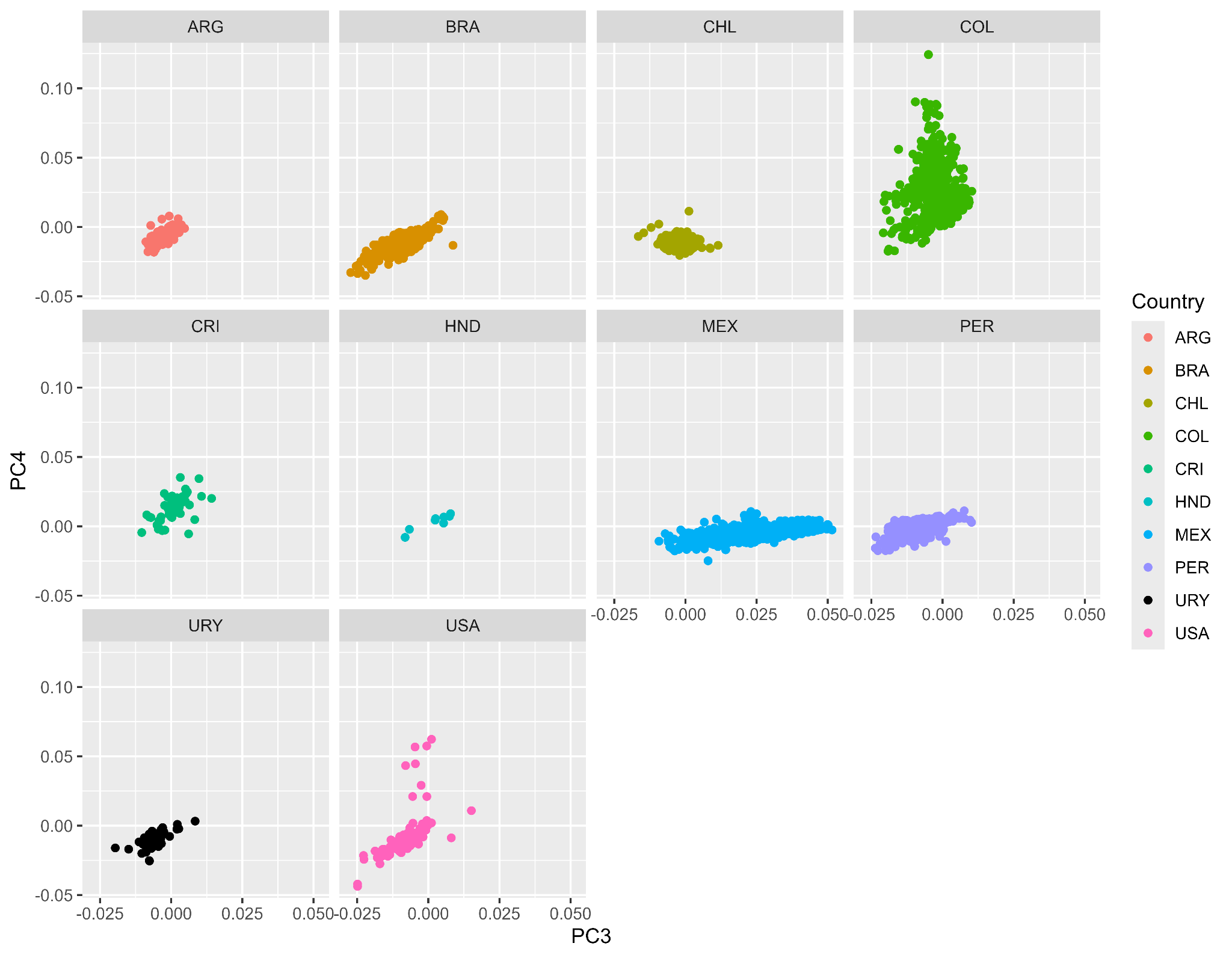

### **Figure S12:** Principal Component Analysis (PCA) for LARGE-PD Phase 2 (PC3 vs. PC4) excluding outliers detected across the first 10 principal components from the projected PCA of LARGE-PD Phase 2. The PCA was performed using all samples and is plotted by country for visual clarity. ARG: Argentina, BRA: Brazil, CHL: Chile, COL: Colombia, CRI: Costa Rica, HND: Honduras, MEX: Mexico, PER: Peru, URY: Uruguay, USA: United States of America

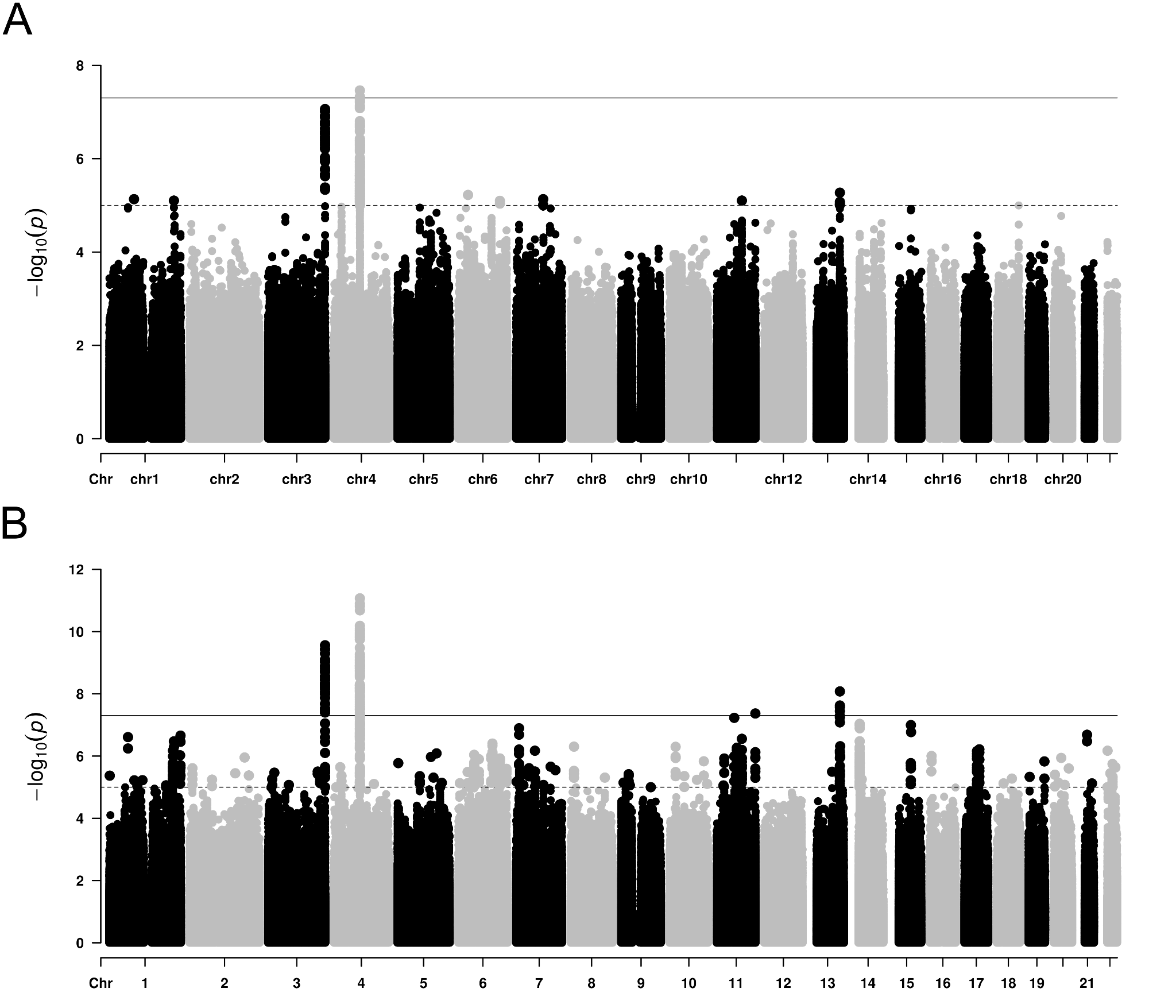

### **Figure S13:** Manhattan plots for (A) SAIGE and (B) ATT for discovery cohort.

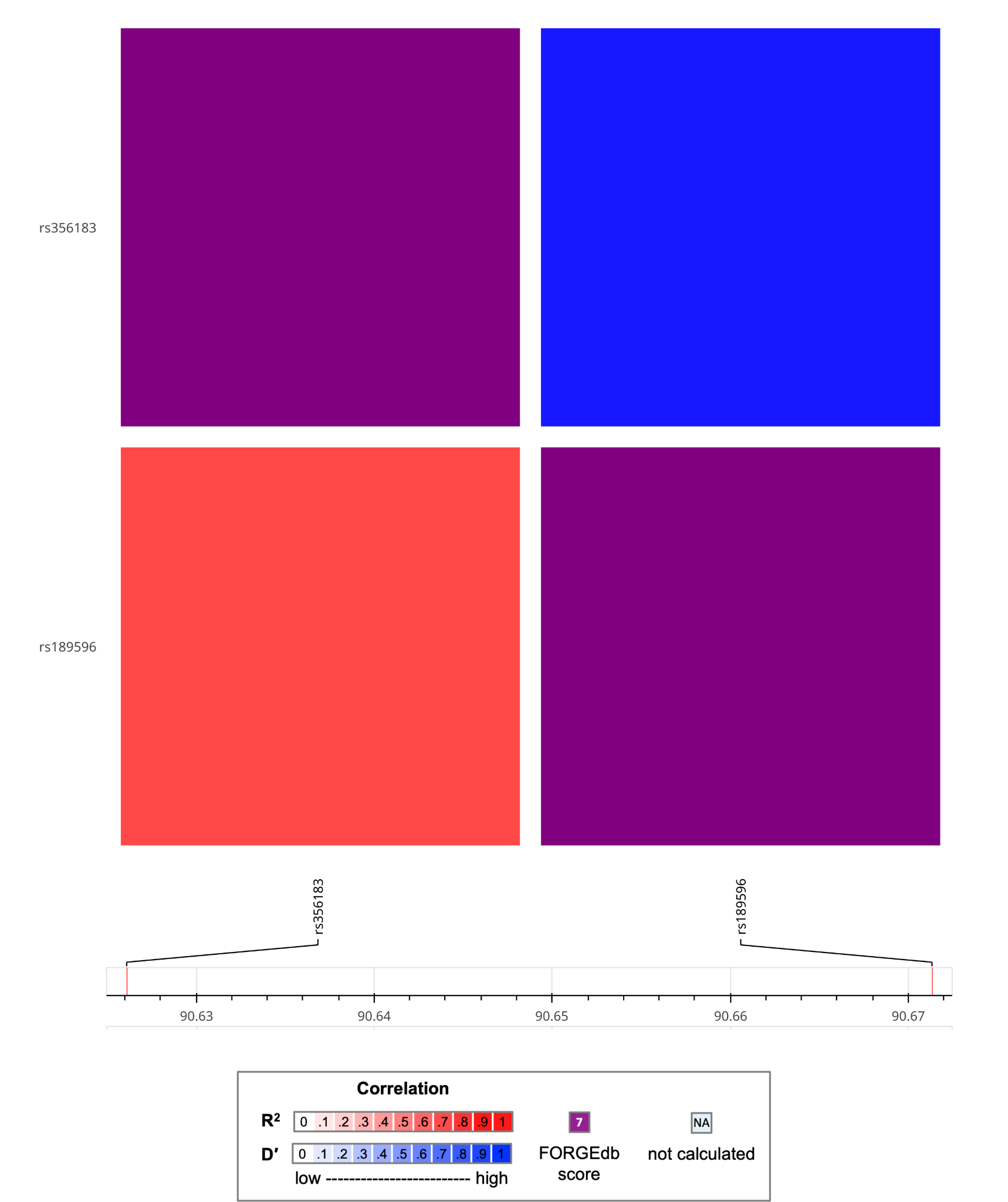

### **Figure S14:** LD Matrix for SAIGE results. In this analysis we used the Admixed American panel as LD reference. Some variants may be not represented because LDMatrix does not accept multi-allelic variants

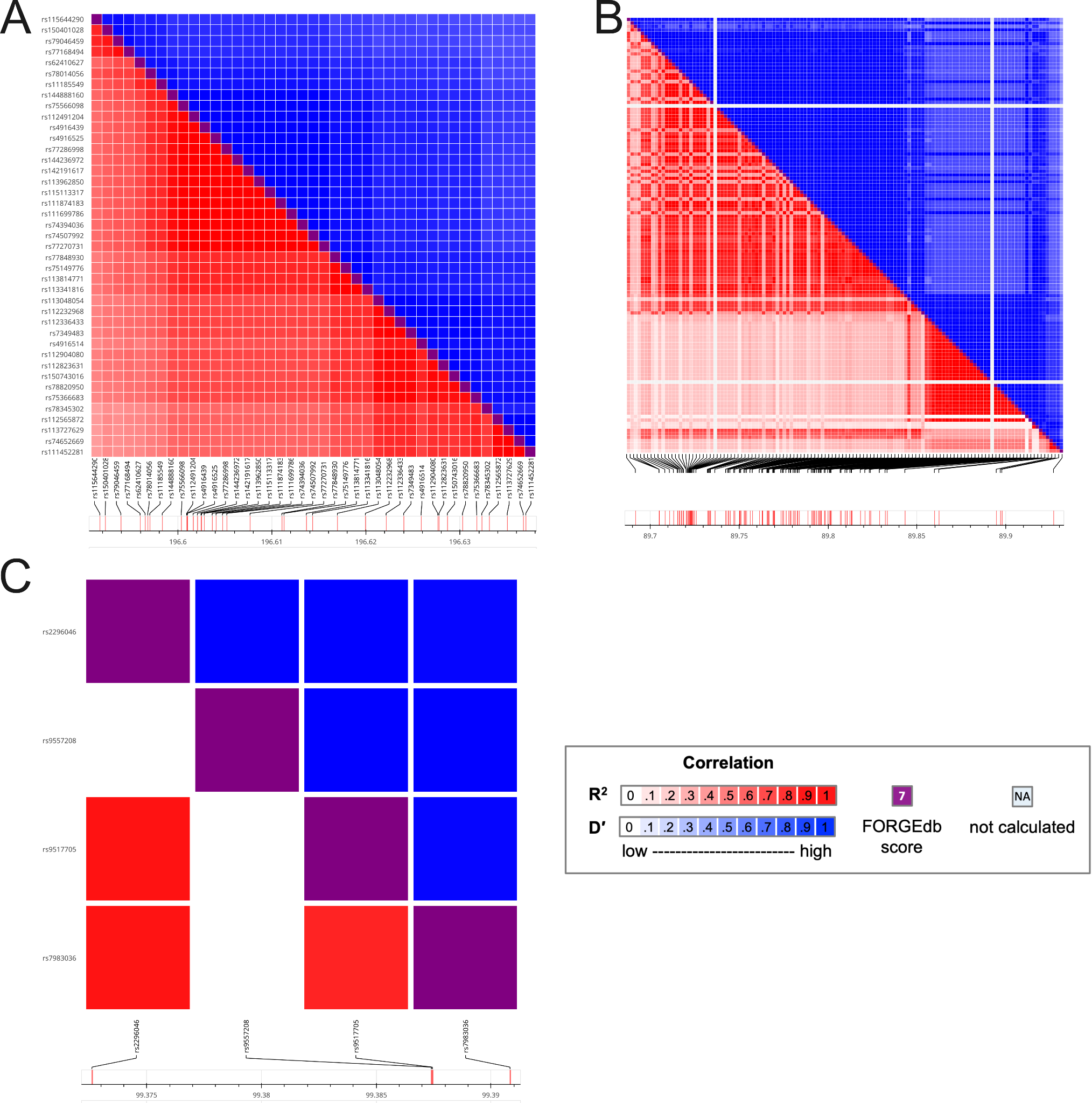

### **Figure S15:** LD Matrix for ATT results for variants on chromosome (A) 3, (B) 4 and (C) 13. In this analysis we used the Admixed American panel as LD reference. Some variants may be not represented because LDMatrix does not accept multi-allelic variants

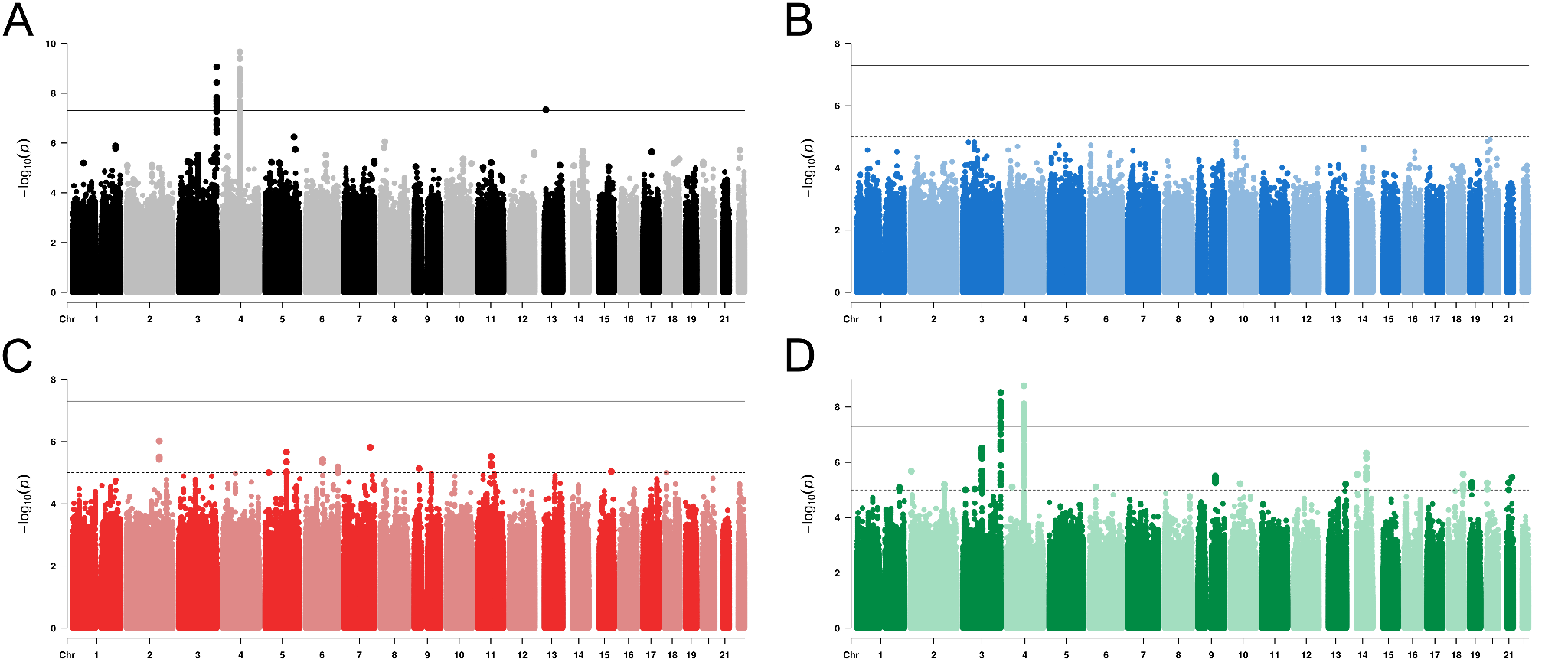

### **Figure S16:** Manhattan plots for TRACTOR results for the discovery cohort. In (A) we have the joint ancestry test and results split by (B) African, (C) European, and (D) Native American ancestries**.**

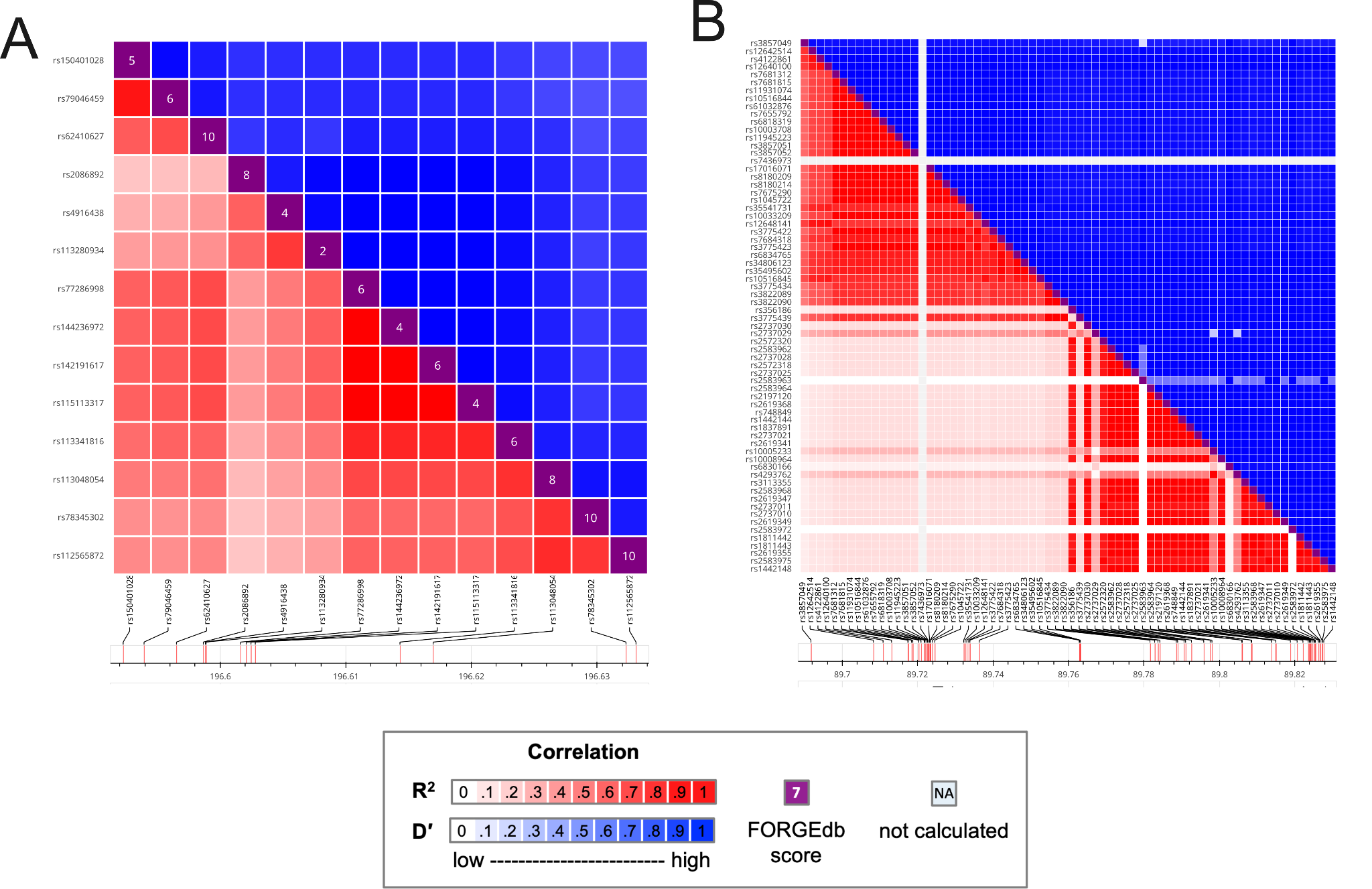

### **Figure S17:** LD Matrix for TRACTOR results for variants on chromosome (A) 3 and (B) 4. In this analysis we used the Admixed American panel as LD reference. Some variants may be not represented because LDMatrix does not accept multi-allelic variants

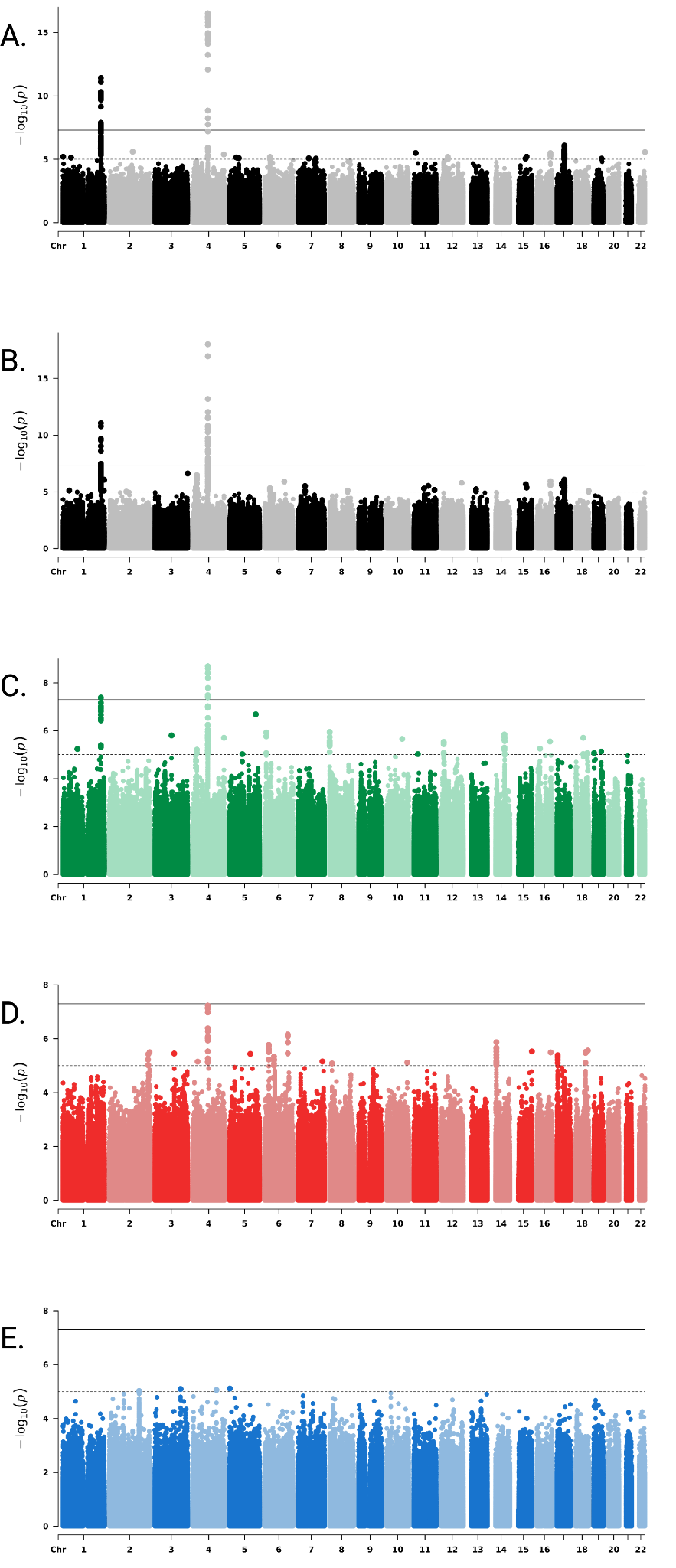

### **Figure S18:** Manhattan plots from random-effects genome-wide association meta-analysis (GWAMA) of LARGE-PD Phase 1 and 2 cohorts for: **A**: SAIGE results meta-analysis. **B**: ATT results meta-analysis. TRACTOR results for Native American (C), European (D), and African (E) ancestry meta-analyses. Each plot shows –log₁₀(p) values across the genome; the horizontal line indicates genome-wide significance of suggestive (1e-5) and statistical significance (5e-8), where notable loci *SNCA* and *ITPKB* are highlighted.

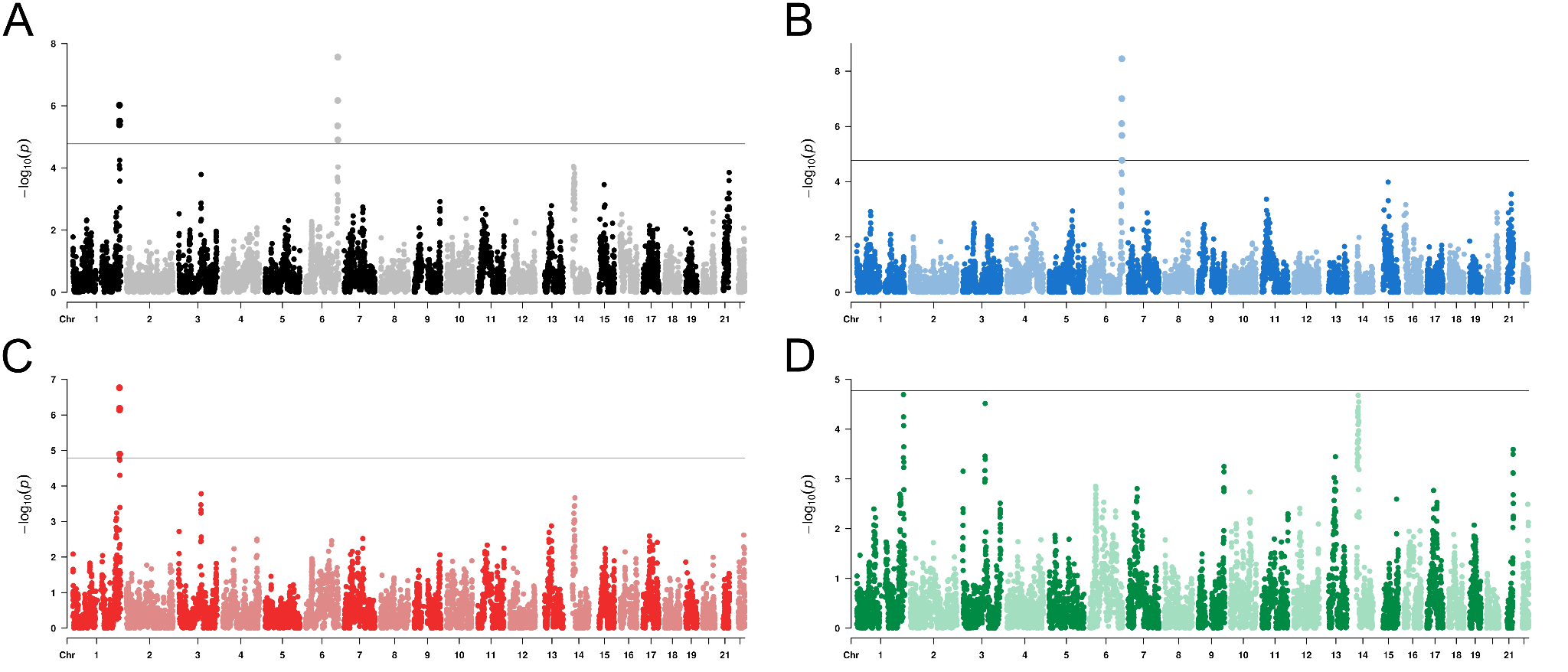

### **Figure S19:** Manhattan plots for Admixture Mapping results. n (A) we have the joint ancestry test and results split by (B) African, (C) European, and (D) Native American ancestries**.**

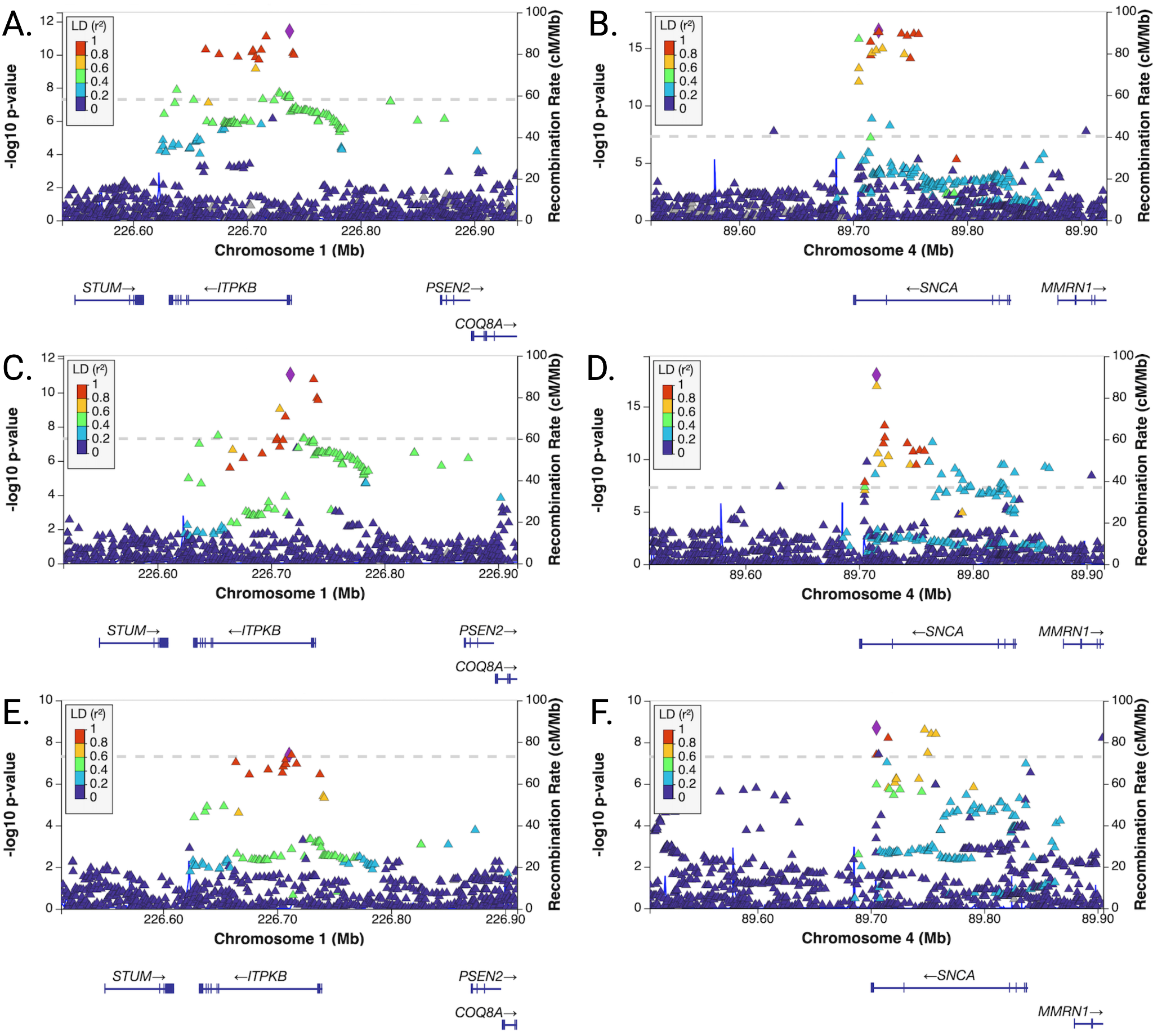

### **Figure S20:** Locus zoom plots for the genomic risk loci identified by **A:** SAIGE meta-analysis on chromosome 1, **B:** SAIGE meta-analysis chromosome on 4, **C:** ATT meta-analysis chromosome on 1, **D:** ATT meta-analysis chromosome on 4, **E:** TRACTOR-NAT meta-analysis on chromosome 1 and **F:** TRACTOR-NAT meta-analysis on chromosome 4.

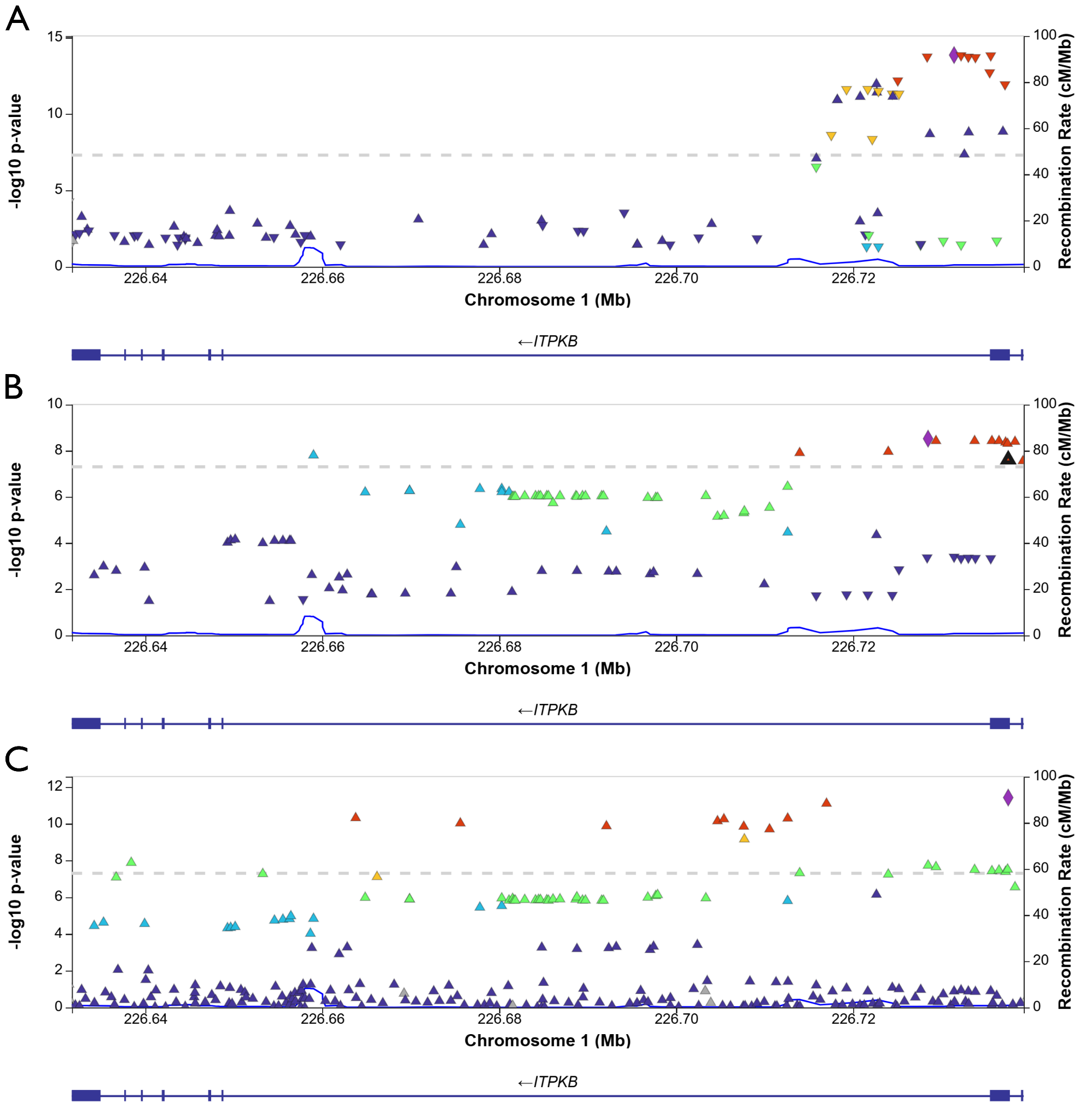

Figure S21: *ITPKB* Locus zoom for (A) European GWAS from The Global Parkinson’s Genetics Program (GP2) et al. 2025, (B) South Asian GWAS from Kishore et al. 2025, and (C) LARGE-PD P1 and P2 meta-analysis. The rs117185933 was not significantly associated on EUR GWAS, but achieved statistical significance on SAS GWAS (p = 2.44 e-08, black triangle) and LARGE-PD.

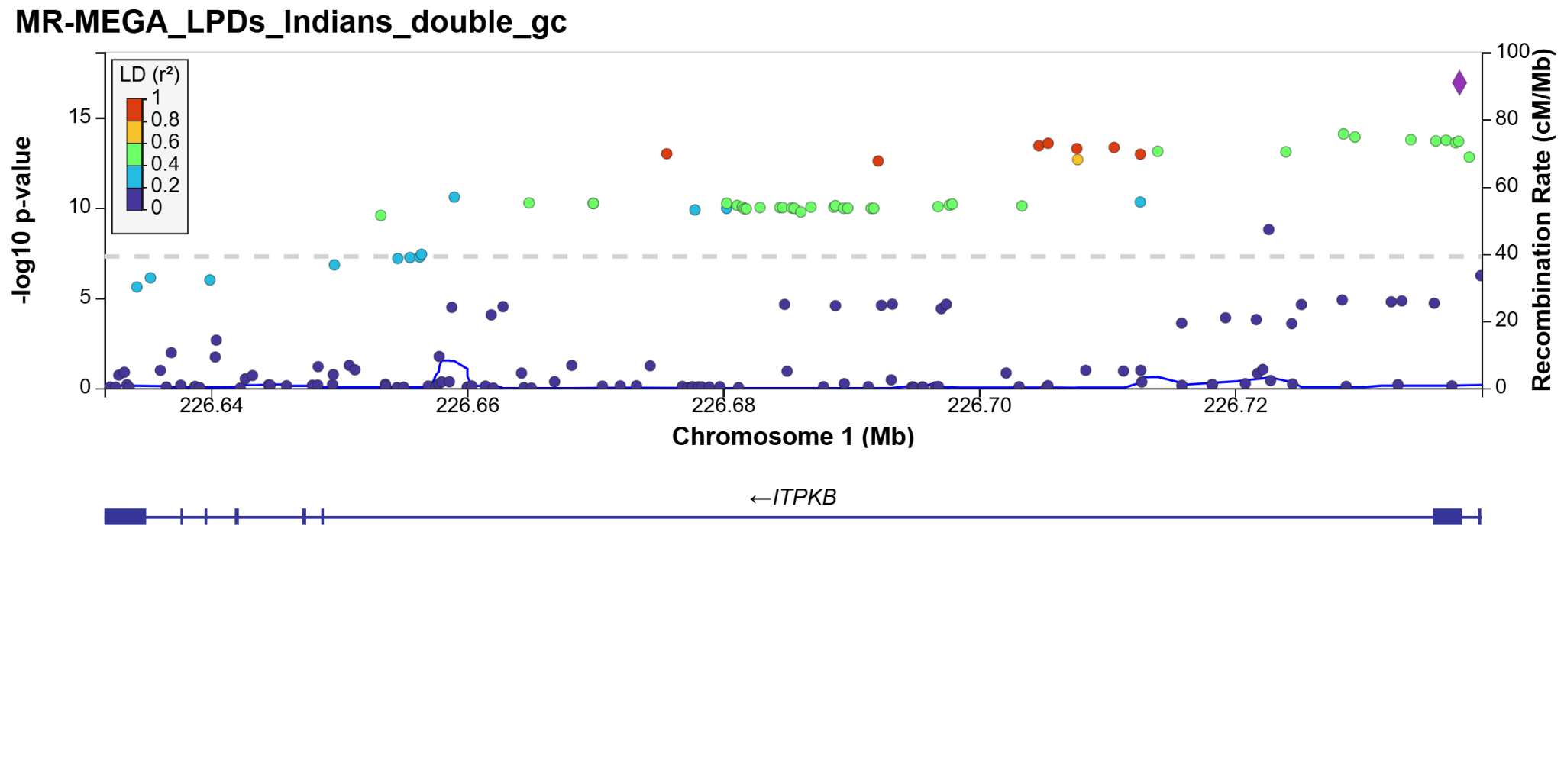

Figure S22: MR-MEGA results for the *ITPKB* gene. This meta-regression analysis was conducted using data from LARGE-PD Phase 1 and Phase 2, Andrews et al. (2024), and the South Asian GWAS by Kishore et al. (2025). In this analysis, the lead variant identified in LARGE-PD emerged as the candidate variant. Colors in the plot represent the linkage disequilibrium (LD) based on the AMR panel from the 1000 Genomes Project.

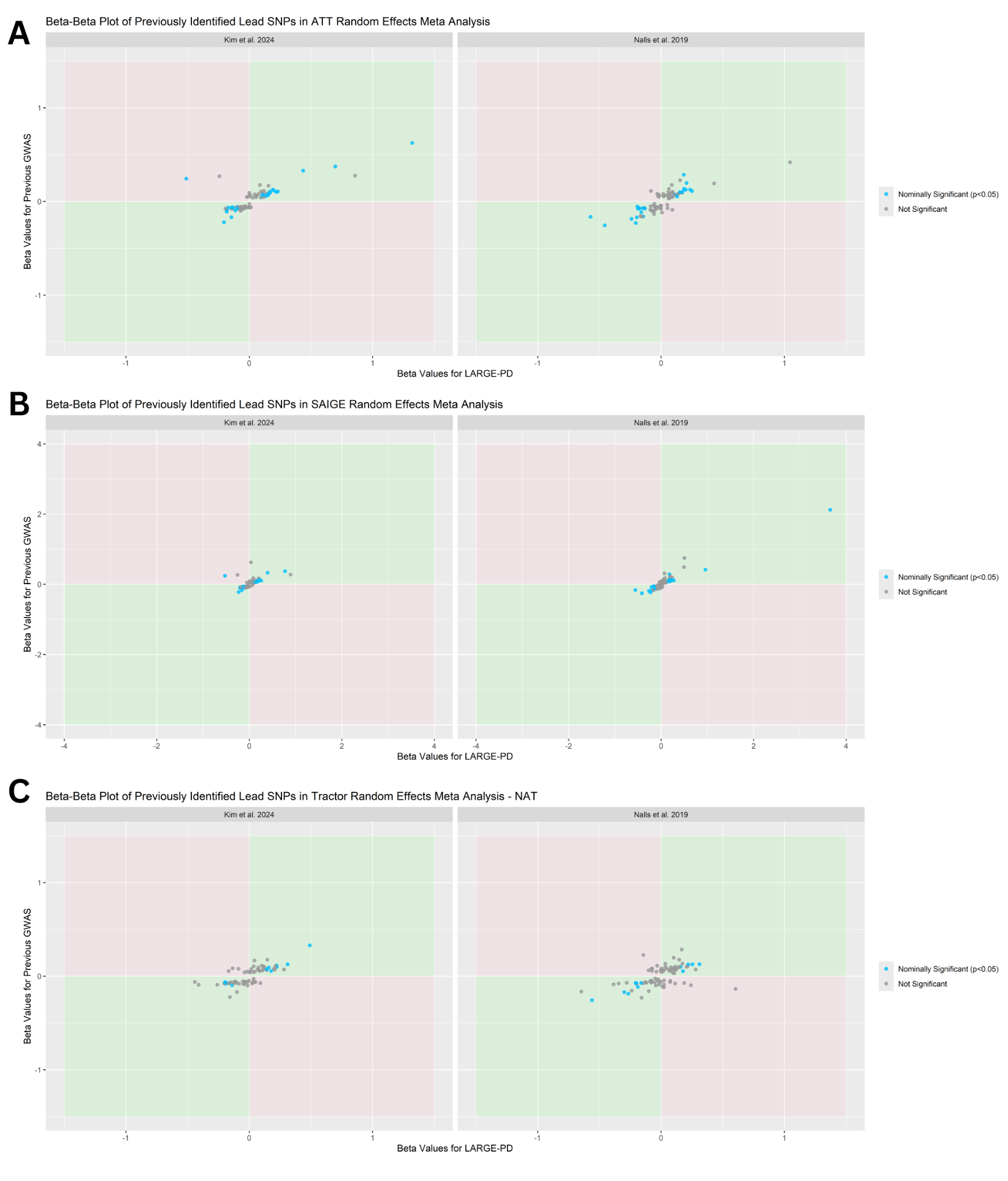

**Figure S23**: Beta-beta plot of previously identified GWAS hits from Kim et al. 2024 and Nalls et al. 2019 compared to the (A) ATT and (B) SAIGE and (C) Tractor-NAT GWAS meta-analyses. Effect sizes and directions from LARGE-PD were compared to Kim et al. 2024 and Nalls et al. 2019. Variants replicated at p<0.05 in LARGE-PD meta-analyses are highlighted. Pearson correlation coefficients for replicated variants: (A) Kim et al. 2024 r=0.83; Nalls et al. 2019 r=0.90, (B) Kim et al. 2024 r=0.69; Nalls et al. 2019 r=0.99, (C) Kim et al. 2024 r=0.97; Nalls et al. 2019 r=0.98.

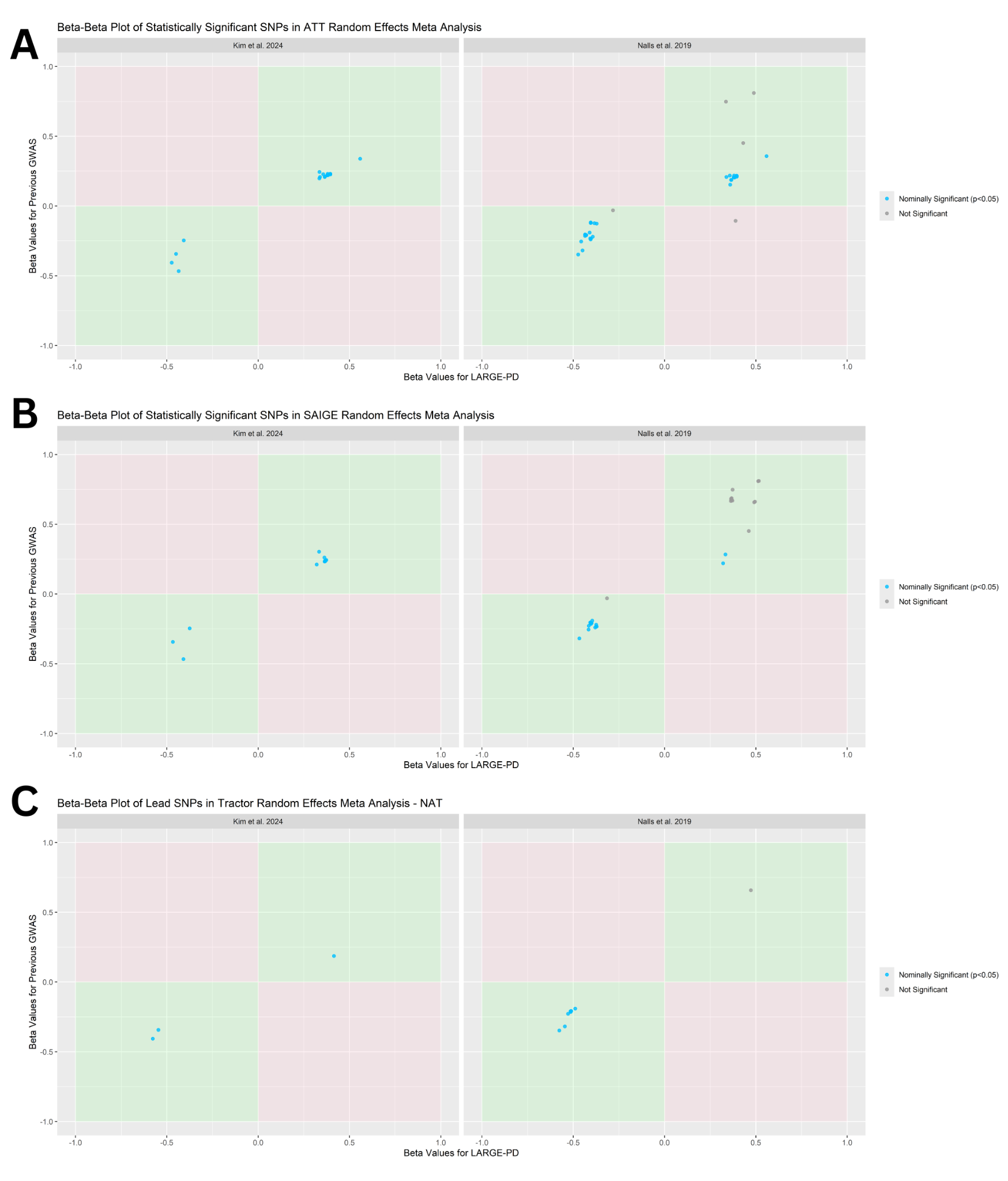

**Figure S24**: Beta-beta plot of suggestive p-value (1x10^-5^) variants from the (A) ATT and (B) SAIGE and (C) Tractor-NAT GWAS meta-analyses compared to previous GWAS. Effect sizes and directions compared to Kim et al. 2024 and Nalls et al. 2019. Variants replicated at p<0.05 in previous studies are highlighted. Pearson correlation coefficients for replicated variants: (A) Kim et al. 2024 r=0.99; Nalls et al. 2019 r=0.98, (B) Kim et al. 2024 r=0.98; Nalls et al. 2019 r=0.99, (C) Kim et al. 2024 r=0.99; Nalls et al. 2019 r=0.95.

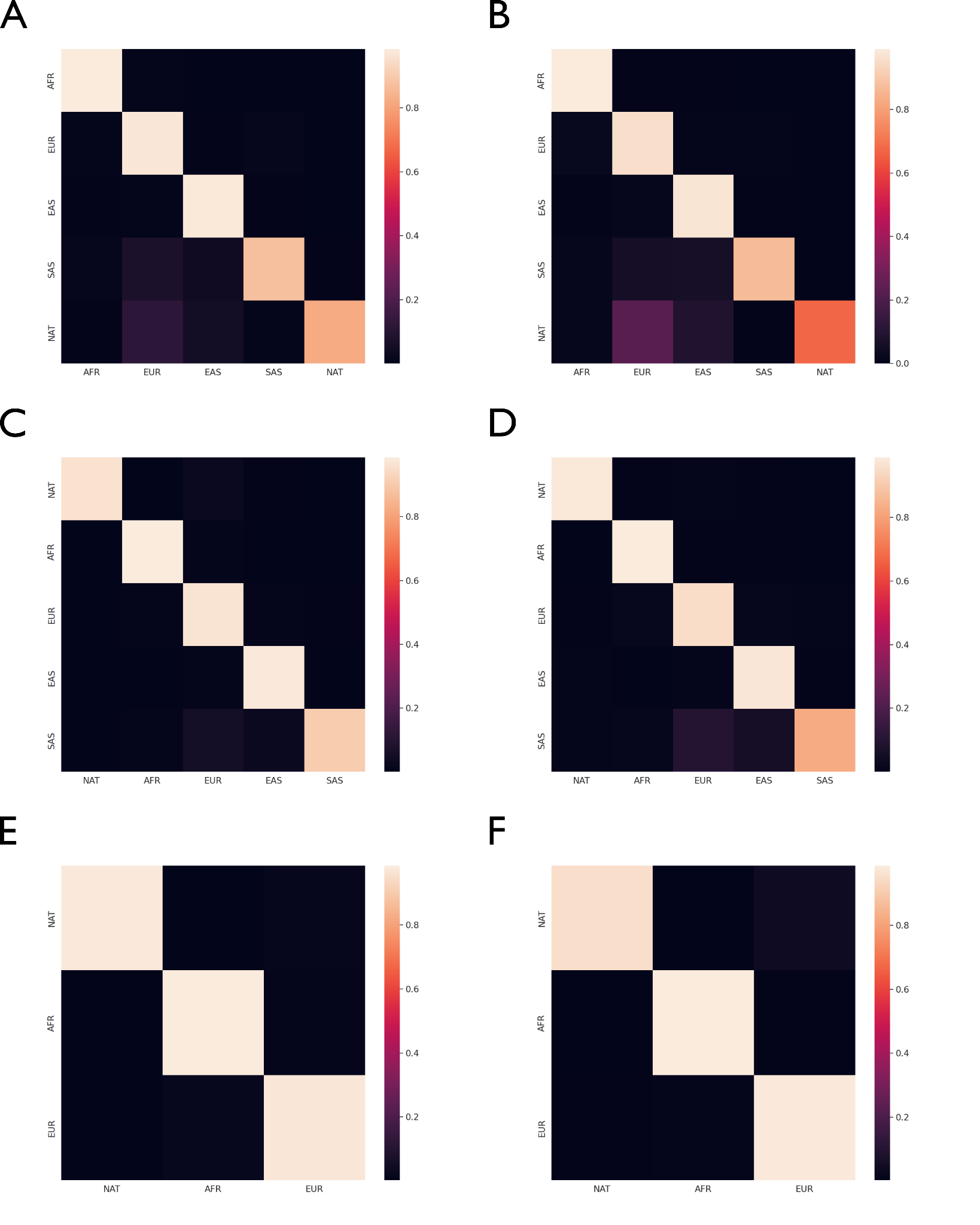

**Figure S25**: Confusion matrices of the validation step provided by G-Nomix. Confusion matrices for a 5-way reference panel using only the 1000 Genomes reference for chromosome 4 (A) and chromosome 17 (B). Confusion matrices for a 5-way reference panel using the 1000 Genomes reference combined with LARGE-PD non-admixed samples for chromosome 4 (C) and chromosome 17 (D). Confusion matrices for a 3-way reference panel using the 1000 Genomes reference combined with LARGE-PD non-admixed samples for chromosome 4 (E) and chromosome 17 (F).

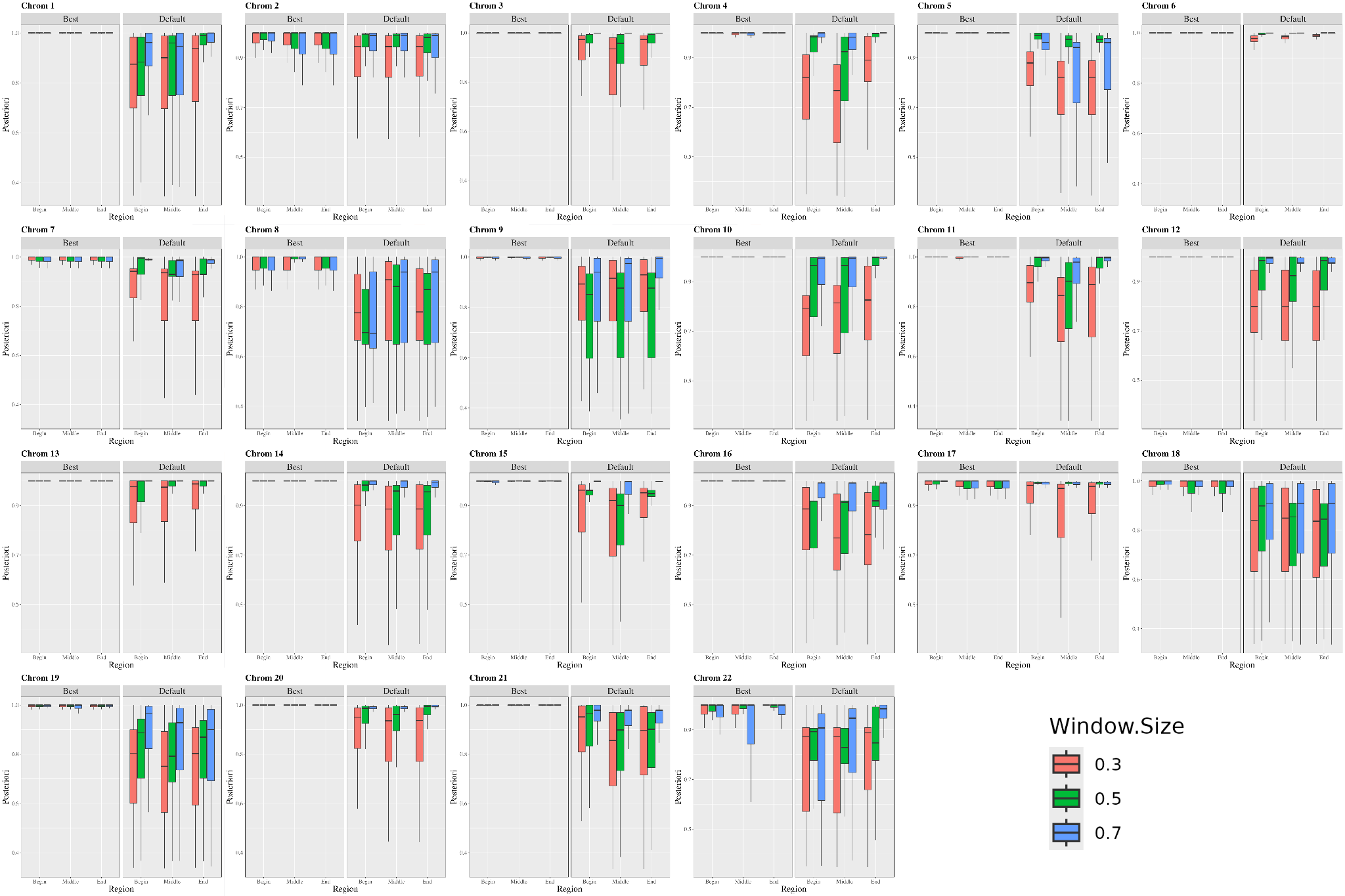

**Figure S26**: Posterior probabilities for all chromosomes using the "default" and "best" models with window sizes of 0.3 cM, 0.5 cM, and 0.7 cM. Posterior probabilities were analyzed across three chromosomal regions—begin, middle, and end—to assess quality based on chromosomal location.
